# Decision models can offload structured clinical tasks from large language models

**DOI:** 10.64898/2026.10.09.26365130

**Authors:** Dean Ariel, Guy Laban, Stav Endelman, Ran Balicer, Noa Dagan, Seffi Cohen

## Abstract

Decision models like Jev read text and return probabilities, categories, or ordinal levels, with confidence scores for the latter two, at a fraction of large language models’ (LLMs) cost. We explored how Jev could be integrated into clinical workflows for all these output types: mortality risk estimation, categorical Choosing-Wisely decisions for patient questions, and ordinal colon cancer staging. With standard LLM-style inputs, Jev trailed five flagship LLMs: mortality AUROC was 0.822 versus 0.837–0.846; Choosing-Wisely accuracy, 87.7% versus 98.4–100%; and staging accuracy, 45.4% versus 94.2–100%. Developer-recommended input alterations improved staging accuracy to 75.8% with medical context, 82.1% with task decomposition, and 85.8% with both. None of Jev’s staging errors had full confidence. Using Jev for fully confident staging decisions and LLMs for the rest was non-inferior to each LLM alone and cut flagship LLM costs by 33–44%. Prompted as designed, decision models may efficiently share structured clinical tasks with LLMs.

## Main text

General-purpose large language models (LLMs) are increasingly used for clinical question answering^1^,^2^. Despite their impressive accuracy, their use at health-system scale can potentially be costly^3^. This motivates exploring ways to reduce this cost.

Decision models such as Jev (TypeSafe AI)^4^ are a new class of AI built for closed-form output questions. They read text but return only a probability, a categorical choice among options, or an ordinal level, the latter two with confidence scores, at a fraction of LLM costs^4^,^5^. Jev’s developers recommend structured questions, preferably with relevant context^5^. These models thus present the opportunity to reduce overall costs, by using them for questions that directly ask for closed-form outputs, or for specific components of the answer-generating process that require such outputs.

We tested how Jev, in its original version released in September 2026, could be integrated into clinical workflows in three steps: with standard LLM-style inputs; with inputs altered to the recommended form; and paired with an LLM (Jev answering when fully confident; the LLM otherwise). These experiments used five flagship and five smaller LLMs.

We explored Jev’s behavior on three clinical tasks, one per output type. For a probability, the task was ten-year mortality-risk estimation for 12,697 adults in the National Health and Nutrition Examination Survey (NHANES)^6,7^. For a categorical choice, we used 180 patient questions asking whether a test or treatment was needed (e.g., a spine MRI for back pain). In 125 questions, the intervention was low value under one of 14 Choosing Wisely recommendations^8^, and in 55, it was indicated (constructed and adjudicated by two physicians). For an ordinal level, the task was colon cancer staging based on the American Joint Committee on Cancer (AJCC) 8th edition^9^, using 240 synthetic pathology reports built from known T, N, and M categories (constructed from input permutations, with a sample adjudicated by a physician). Each task type was also repeated on a second dataset: in-hospital death probability in 1,297 stays from the open eICU intensive care demo^10,11^, 1,571 categorical board questions from MedQA and Medbullets in the MedHELM benchmark^12^, and 561 cases requiring ordinal staging of acute kidney injury under the ‘Kidney Disease: Improving Global Outcomes’ (KDIGO) criteria^13^.

First, Jev received standard LLM-style inputs (Methods). In this form, it was less accurate than flagship LLMs across all tasks, but was more accurate than some smaller LLMs. For ten-year mortality, AUROC was 0.822 (95% CI 0.813–0.832), compared with 0.837–0.846 for the flagship LLMs and 0.854 for a logistic regression model fitted to the outcomes (Fig. 1a). It was poorly calibrated, with a mean predicted risk of 26.3% whereas 15.8% experienced the outcome (LLMs 18.5–25.2%, Supplementary Table 1). In intensive care, Jev’s hospital mortality AUROC was 0.836 versus 0.845–0.880 for flagship LLMs. On Choosing Wisely-related patient questions, Jev had a balanced accuracy of 87.7% (95% CI 82.7–92.1%), counting low-value and indicated interventions equally, against 98.4–100.0% for the LLMs (Fig. 1b). Board-question accuracy was 85.6% (LLMs 91.2–95.9%). On cancer staging, it gave the exact stage for 45.4% of the 240 reports, compared to 94.2–100.0% for the LLMs (Fig. 1c). Its errors mattered clinically: Jev placed 9 of 63 stage III reports in stage 0–II, across the line that decides adjuvant chemotherapy. Kidney-injury staging accuracy was 54.4% versus 90.9–99.5% for LLMs. Jev was also faster and cheaper: it answered in a median of 0.90–1.02 seconds, compared with 2.5–36.8 seconds for the LLMs, and cost US$0.02–0.04 per 1,000 answers, compared with US$1.56–15.08 (Fig. 1d). Further results appear in Supplementary Tables 2–4.

**Figure 1.**
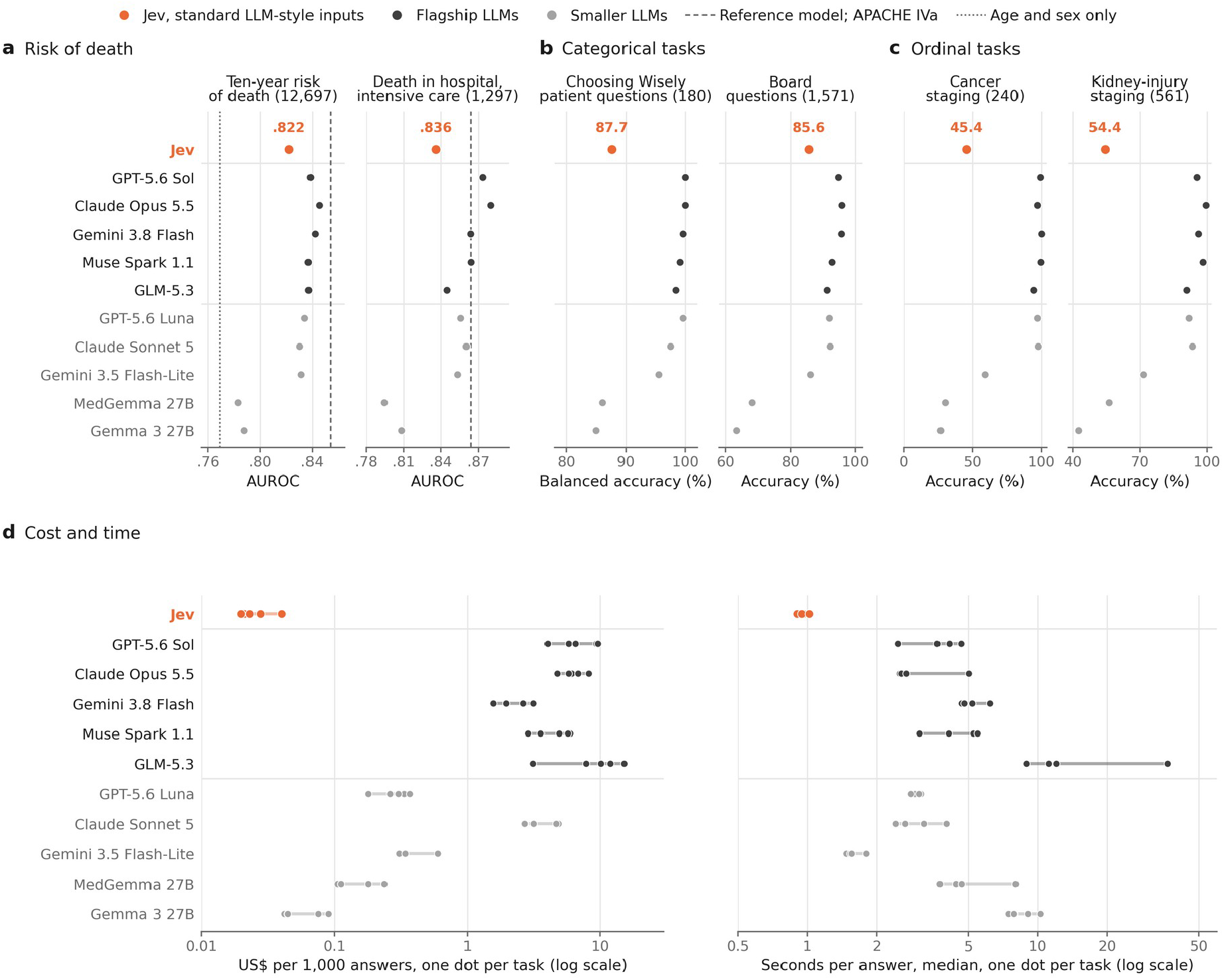
Jev’s standalone performance compared to LLMs, given standard LLM-style inputs. **a**, Risk of death, AUROC: ten-year risk in NHANES adults, with a reference model (dashed) and age and sex only (dotted); death in hospital in intensive care, with APACHE IVa (dashed). **b**, Categorical tasks: balanced accuracy for Choosing Wisely-related patient questions and accuracy for board questions. **c**, Ordinal tasks: cancer staging and kidney-injury staging. **d**, Cost per 1,000 answers (five tasks) and median time per answer (four timed tasks), one dot per task; log scales. 95% CIs and costs: Supplementary Tables 1–4.

Next, we tested two developer-recommended input alterations^5^: structuring and added medical context. Input structuring turns numbers into named categories and splits a broad judgment into simple questions whose answers are combined into a final answer by deterministic code (task decomposition). Medical context provides definitions for each answer option. For the risk-of-death estimations, each numeric value in the record was replaced by its clinical category, so an HbA1c of 8% became “diabetes range” and a creatinine level became a kidney function stage. The categorical tasks, which already consisted of clearly defined options, remained unchanged. For cancer staging, we tested the two types of alterations. Medical context added each stage’s T, N, and M categories. For task decomposition, we broke the task into four questions (how deeply the tumour had grown, how many regional nodes were affected, whether tumour deposits were present, and whether it had spread to distant sites), and deterministic code looked up the stage in the AJCC table. For kidney-injury staging, we added each stage’s KDIGO criteria for context.

For mortality estimation, these changes left AUROC unchanged and calibration poor (Supplementary Table 1). On cancer staging, each alteration improved Jev’s accuracy (Fig. 2a and Supplementary Table 5): medical context alone raised it from 45.4% to 75.8%, task decomposition alone to 82.1%, and both to 85.8%. Flagship LLM performance changed little, from 94.2–100.0% to 98.8–100.0% with both changes. Adding each stage’s KDIGO criteria raised kidney-injury staging accuracy from 54.4% to 91.8% for Jev and from 90.9–99.5% to 97.9–100.0% for the LLMs.

**Figure 2.**
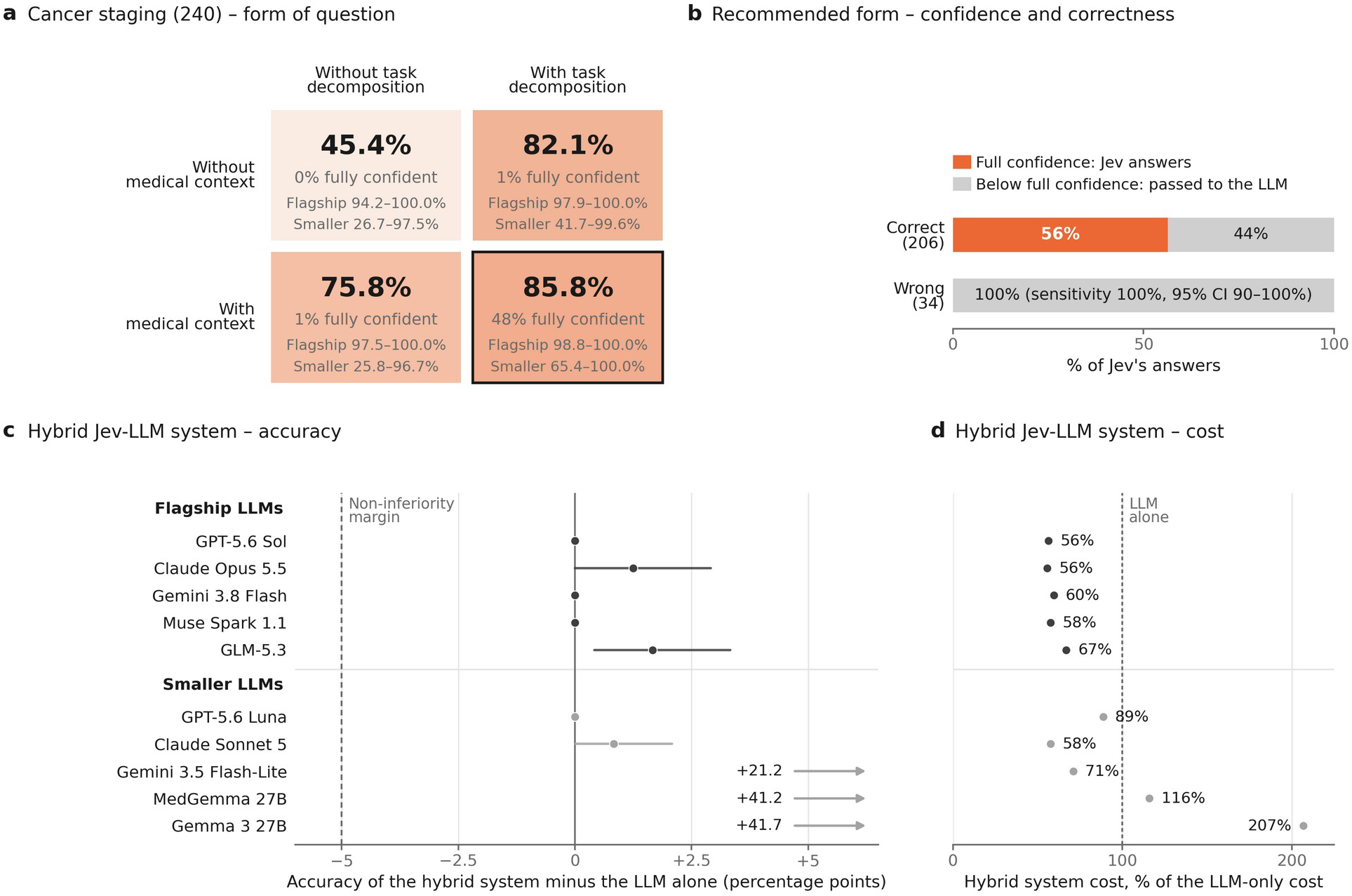
Performance changes with developer-recommended input alterations and overall performance of Jev-LLM hybrid systems. Colon cancer staging on 240 pathology reports. **a**, Jev’s staging accuracy by form of question, along with the share of its answers that received full confidence (1.00), and the accuracy ranges of the flagship and smaller LLMs given the same form. **b**, Jev in the recommended form: breakdown of correct and wrong answers by whether they reached full confidence. **c**, Hybrid Jev-LLM system performance compared to the LLM alone, with Jev answering when fully confident and each of ten LLMs otherwise (dark grey, flagship; light grey, smaller; 95% bootstrap intervals). Dashed line: 5-point non-inferiority margin. Arrows mark differences beyond the axis. **d**, Hybrid cost relative to LLM alone. Additional results by report type: Supplementary Tables 5 and 7a.

Jev’s confidence in its chosen answer depended on question form (Supplementary Table 6). Given standard LLM-style inputs, Jev was never fully confident in either staging task, and assigned confidence below 0.75 to 98% of cancer-staging answers. With recommended inputs, it was fully confident on 48% of pathology reports, which were all indeed correct. Across all tasks and forms, 99.8% (1,351 of 1,354) of Jev’s fully confident answers were correct. When the flagship LLMs were also asked to assign confidence to their answers, although this is not a native feature as in Jev’s case, they gave 78–97% of their answers, correct or wrong, a confidence between 0.75 and 0.99, and were rarely fully confident (0–21%). This pattern also persisted when the inputs were entered with the additional information recommended for Jev’s operation (0–25% of reports, none wrong). Three smaller LLMs also gave fully confident incorrect answers (Gemini 3.5 Flash-Lite in 29 of the 240 reports, MedGemma 27B in 46, and Gemma 3 27B in one).

Finally, we paired Jev with an LLM in a hybrid system to balance accuracy and cost. For cancer staging with recommended inputs, Jev answered only at full confidence (1.00 on all four questions), passing other reports to an LLM. None of Jev’s 34 errors on the 240 pathology reports was fully confident; 56% of its correct answers were fully confident (Fig. 2b and Supplementary Table 6). In this Jev-LLM pairing, Jev correctly answered 116 of 240 reports (48.3%) itself. The pairing with every flagship LLM was at least as accurate as the LLM alone (95.8–100.0%, differences 0.0 to 1.7 points; Fig. 2c). All ten pairings were non-inferior within a 5-point margin. With flagship LLMs, hybrid staging cost 56–67% as much as the LLM alone and took an estimated 55–70% of the time (Fig. 2d and Supplementary Tables 4 and 7). The three LLMs with the poorest initial accuracy gained the most: paired with Jev, they were 21–42 points more accurate in cancer staging (Fig. 2c). These inexpensive models yielded smaller or no cost savings, and two pairings increased costs (Fig. 2d). Similar patterns repeated for other tasks. Jev answered 55% of patient questions, 30% of board questions, and 7% of kidney-injury stagings; every pairing remained non-inferior to its LLM alone. For flagship LLMs, savings tracked the share Jev answered: about half the cost for patient questions and little for kidney injury (Supplementary Table 7b).

In summary, Jev is not designed to replace LLMs on every clinical task, and its performance with standard LLM-style inputs indeed did not match that of flagship LLMs. However, for suitable tasks, particularly structured outputs based on defined clinical rules, and following the developers’ recommended input format, it can produce meaningful outputs. Most strikingly, its fully confident answers were almost always (99.8%) correct in our experiments, making confidence a practical signal for dividing work between Jev and an LLM. Hybrid systems built on this principle were non-inferior to LLMs alone in terms of accuracy and achieved substantial cost reductions. Our results provide preliminary evidence of the efficiency of these hybrid systems, but they require further evaluation in real-world clinical tasks reflecting the complexity of real patients. Nonetheless, our findings suggest a promising new role for decision models: offloading structured clinical tasks from LLMs to reduce cost and response time.

## Methods

We compared Jev with widely used LLMs on three kinds of clinical questions, one per type of output: a probability (ten-year risk of death; death in hospital in intensive care), a categorical choice (patient questions; board questions), and an ordinal level (cancer staging; kidney-injury staging). Within a task, every system received the same cases, and answers were scored against keys that depended on no model or rater: observed deaths, the guideline or published answer, and the AJCC or KDIGO tables. The patient questions and pathology reports were created for this study and are unpublished.

### Systems

#### Jev

Jev 1.13 (TypeSafe AI) answers a typed question about a case given as text^4,5^: a Noul question returns the probability that a statement is true, a Choice question a probability for each option, and a Score question a probability for each level of an ordered scale^5^.

#### LLMs

The five flagship LLMs were GPT-5.6 Sol (OpenAI)^14^, Claude Opus 5.5 (Anthropic)^15^, Gemini 3.8 Flash (Google)^16^, Muse Spark 1.1 (Meta)^17^, and GLM-5.3 (Z.ai)^18^, each in a version available in September 2026. The five smaller LLMs were the models in the free OpenAI, Anthropic, and Google plans, GPT-5.6 Luna^14,19^, Claude Sonnet 5^20^, and Gemini 3.5 Flash-Lite^21^, with MedGemma 27B, an open model trained for medicine^22^, and Gemma 3 27B, from which it was built^23^ (Supplementary Table 8).

#### Settings and prompts

Every system ran at its provider’s default settings, with output capped at 16,000 tokens and the seed 20260922 where accepted, through OpenRouter^24^ to one first-party provider per model with fallbacks disabled; MedGemma and Gemma went through the Hugging Face router. Calls were made on 26–30 September 2026, and the first stored response to each request was analyzed. Within a task, Jev and the LLMs received character-identical cases, questions, and options, and every request followed one template in which only the case changed. The LLMs also received a system message specifying the reply format; replies were constrained to a JSON schema that was supported and read by a single parser for each system (Supplementary Note 1).

#### Recommended form

Jev’s documentation recommends named fields, numbers given as named categories, each option defined by what it includes and excludes and by examples, arithmetic done in code, and a broad judgment split into simple questions that code combines^5^. We applied this wherever it changed the question: for risk of death, numbers became categories, and for cancer staging, the staging rule became four questions (Supplementary Note 2). The patient questions and board questions were already one choice among defined options. For kidney-injury staging, the recommended form was the version with each stage’s KDIGO criteria, because splitting the rule further would have left only arithmetic. For risk of death, only Jev was called again. For cancer staging, every LLM answered every form Jev answered, with the same reports, questions, and options, and its stage was computed from its four answers in the same way (Supplementary Note 1).

### Ten-year risk of death (probability)

#### Data

NHANES 1999–2008 was linked to the public-use Linked Mortality File, with deaths through 31 December 2019^6,7^. Participants aged 40–79 and eligible for linkage were included; the outcome was death from any cause within 120 months of examination, and participants whose vital status at 120 months was undetermined were excluded (Supplementary Table 9). Participants with all nine of a set of routine blood biomarkers were randomly assigned (seed 20260922) to an evaluation set (1,000), a software-test set (200), and a fitting set; those missing one or two joined the fitting set only.

#### Question

Each record listed named fields with values and units (Supplementary Note 1). Every system was asked about the statement “This person will die from any cause within ten years of this examination.” Jev returned its probability; the LLMs, given a system message describing a mortality-risk study, returned a probability between 0 and 1.

#### Reference and measures

The reference was a ridge-penalised logistic regression of ten-year death on every recorded variable, with cubic splines for continuous variables and missingness indicators, fitted on the fitting set (on 1,200 held-out participants, AUROC 0.826, calibration slope 0.834; Supplementary Note 3). Prediction was scored on the 12,910 records outside the software-test set with all nine biomarkers (the evaluation set and the complete records of the fitting set), the reference contributing ten-fold out-of-fold predictions for the fitting set, and systems were compared on the 12,697 records every system answered: AUROC (95% bootstrap interval, 2,000 resamples), mean predicted risk against the observed share who died, calibration slope, and log-loss, with probabilities clipped to [0.005, 0.995] (Supplementary Table 1). Jev was non-inferior to an LLM when the upper simultaneous 95% bound of the log-loss difference lay below 10% of the gap between a model of age and sex only and the reference. How estimates changed when one field of a record was edited is reported in Supplementary Table 10, and a masked-field recovery test screened for prior exposure to the records (Supplementary Table 11).

#### Recommended form

Each numeric field was replaced by the name of its published clinical category, for example, WHO body-mass-index classes and KDIGO G1–G5 from the race-free CKD-EPI 2021 equation; categorical fields and missing values were unchanged (Supplementary Table 12).

#### Intensive care

Prediction was repeated on the open-access eICU Collaborative Research Database demo (version 2.0.1)^10,11,25^: 1,297 first intensive care stays of adults with an APACHE IVa result who were alive 24 hours after admission, each described by its worst first-day values, with the statement “This patient will die before hospital discharge.” Jev and all ten LLMs answered each stay once. Reference models (spline logistic regression, LightGBM^26^, and XGBoost^27^) were fitted locally on the full credentialed database (version 2.0)^10,28^ without the demo patients’ stays (107,032 stays); only demo records were sent to the models. The predicted mortality of APACHE IVa^10,29^ served as the clinical benchmark. In the recommended form, fields shared with NHANES retained their categories; other laboratory values used the same reference ranges; and fields recorded only in intensive care used the Sepsis-3 categories (SOFA, SIRS, and lactate)^30^ (Supplementary Table 12).

### Patient questions (categorical)

#### Questions

**The authors wrote patient questions for 14 Choosing Wisely recommendations**^**8**^ **on which guidelines agree (Supplementary Table 2c)**, ten per recommendation in which the test or treatment is low-value and five in which it is indicated. Questions that a second physician author did not confirm were excluded, leaving 180 (125 low-value, 55 indicated).

#### Question and measures

Each recommendation had one set of four answers: the test or treatment, holding off with an alternative plan, and two actions wrong in either kind of question, shuffled once per item and identical for every system. Holding off was correct when the test or treatment was low-value, and the test or treatment when it was indicated. The primary measure was balanced accuracy, the mean of the accuracies in the two kinds of question, with overuse and false restraint also reported. Unparsable answers counted as wrong. Because the questions of one recommendation share an answer set, comparisons were repeated by resampling whole recommendations (Supplementary Table 2).

#### Board questions

The same systems answered 1,571 board-examination-style questions, the United States test set of MedQA (1,273 questions, four options) and Medbullets (308 questions, five options; ten verbatim repeats counted once), loaded as in MedHELM’s code^12,31,32^, with the same prompt form, parser, and scoring. Each question was also asked in its other published version, with five options for MedQA and four for Medbullets. These questions are public and may have been seen in training (Supplementary Table 2).

### Cancer staging (ordinal)

#### Reports

Public colorectal pathology reports mostly state the T and N categories in the text, under earlier AJCC editions (by our count, 88% of 544 reports in The Cancer Genome Atlas^33^), so we built reports from specified categories, with the 8th edition table as the key. In September 2026, after every stated training cutoff (Supplementary Table 8), so that no model could have seen them, we built a corpus of 240 pathology reports of colon adenocarcinoma resections, half built to mislead, in a synoptic format adapted from the College of American Pathologists protocol^34^ or as narratives. Each report was built from T, N, and M categories drawn uniformly among the combinations of its stage group, except that Tis occurred only in stage 0 and a stage IVC report that drew T1 or T2 had T redrawn from T3–T4b. Node counts fell within the N category (at least 12 examined), tumour deposits appeared in every N1c report and in 25% of other node-positive reports, and M was stated in a line on staging computed tomography. No report named a category or a stage (Supplementary Note 4). Only colon tumours were used, because pathological T4a is ambiguous in the rectum.

#### Key

The stage group followed from T, N, and M through the AJCC 8th edition table^9^, checked against a grid entered separately. The scale had ten levels (0, I, IIA, IIB, IIC, IIIA, IIIB, IIIC, IVA–IVB, and IVC); IVA and IVB shared a level because a Score question takes at most ten^5^. Every report was checked against its intended T, N, and M (Supplementary Note 4).

#### Question

Each system was asked to “Assign the pathological stage group for this colorectal cancer according to the AJCC Cancer Staging Manual, 8th edition. Stage IVA-IVB here includes IVA and IVB.” once with the stage names only and once with each stage’s T, N, and M categories (medical context). Jev’s answer was its most probable level; the LLMs returned a stage and a probability for each level. The primary measure was exact accuracy with stage names (Supplementary Table 3).

#### Recommended form

The report was sent with four choice questions: the T category, the number of involved regional nodes (0.2 mm or more), tumour deposits, and distant metastasis. Each option included its AJCC definition, what it excludes, and two examples not found in any report. The stage group was computed from the four answers with the table that built the key, and Jev’s confidence was the product of the four top probabilities; a product of 1.00 was full confidence. A second version added AJCC notes on isolated tumour cells, adhesions without tumour, and non-regional nodes, whose examples of non-regional nodes named the two sites used in the misleading reports described below; a third sent the four questions with bare option labels (task decomposition only). In every version with the four questions, the report was sent as a named field, as the documentation recommends. With the single-stage question, this did not change accuracy (Supplementary Table 5).

#### Reports built to mislead

One hundred reports, 20 per feature, contained a potentially misleading feature: isolated tumour cells in a node, a tumour adherent to an organ without tumour in the adhesion, perforation through the tumour, metastasis in a non-regional node, and features that do not change the stage. Twenty more placed the non-regional node at a site named in no question, note, or example, and the regional node at a site with no colon segment^34^, to test the notes beyond the sites they name. Because a report in practice does not indicate whether it is hard, results are reported for the whole corpus and by report type in Supplementary Tables 5 and 7a.

### Kidney-injury staging (ordinal)

#### Records and key

eICU demo stays with at least two serum creatinine values were eligible. Each record provided age, sex, admission weight, all creatinine values from 7 days before to 72 hours after unit admission with their times, and any start of dialysis within 72 hours. The key was the highest KDIGO creatinine stage^13^ in the first 72 hours, computed on the values shown: a rise of at least 0.3 mg/dL within 48 hours or to 1.5–1.9 times the lowest value within 7 days gave stage 1, 2.0–2.9 times stage 2, and 3.0 times or more, dialysis, or a value of at least 4.0 mg/dL reached by a rise meeting either criterion gave stage 3. Urine output was not used. Patients on maintenance dialysis, for whom the KDIGO definition is not intended, were excluded. Up to 250 stays per stage were randomly drawn (seed 20260922), one per patient, giving 561 records (250 without kidney injury, 228 at stage 1, 33 at stage 2, and 50 at stage 3), a sample enriched for kidney injury.

#### Question

Each system was asked, “By the KDIGO creatinine criteria, what was this patient’s highest AKI stage during the first 72 hours in the intensive care unit?” once with the stage names only and once with each stage’s KDIGO criteria, and scored by exact accuracy (Supplementary Table 3).

### Confidence and pairing with an LLM

For a categorical choice or an ordinal level, a system’s confidence was the highest probability it gave any option; probability lists summing to 0.9–1.1 were rescaled, and others were left out of confidence analyses. Full confidence in Jev’s recommended form was scored as a test for its errors on the 240 reports and by report type: sensitivity was the share of wrong answers short of it, and specificity the share of correct answers reaching it (Supplementary Tables 6 and 7a). Answers were grouped by confidence: below 0.75, 0.75–0.99, and 1.00 (fully confident). Clinical decisions (ordering the test or treatment, stage III–IV, and stage 2–3 kidney injury) were also scored by sensitivity and specificity (Supplementary Table 13).

In each pairing, built from the stored answers, Jev answered first, and an LLM answered the items Jev passed on; cost counted Jev on every item and the LLM on those passed on. For cancer staging in the recommended form, Jev kept the reports it answered at full confidence, and the LLM answered the rest with stage names or, in a second pairing, in the recommended form; a final answer in a different group of stage 0–II, III, or IV from the key counted as crossing a treatment line. Each of the ten LLMs was paired in this way. The same rule was applied to patient questions, board questions, and kidney-injury staging (Supplementary Table 7b). Lower confidence thresholds were examined descriptively with the same non-inferiority test (Supplementary Table 7c and Supplementary Fig. 1). Risk of death was not included because its answer is a probability of death, which carries no separate confidence and cannot be scored correct or wrong for one person.

### Cost and speed

Cost was calculated from the tokens for each answer at the model’s standard list price in September 2026 (Supplementary Table 8). Savings in US dollars at each confidence threshold are in Supplementary Table 7c. Response time was measured on a separate sample of 100 items per system for risk of death, patient questions, board questions, and cancer staging, sent one at a time, with all systems of a task timed in the same session (Supplementary Table 4). Pairing time was estimated as Jev’s time for reports it kept and the sum of Jev’s and the LLM’s times for reports passed on.

## Data Availability

NHANES and the Linked Mortality Files are available from the National Center for Health Statistics, the eICU demo from PhysioNet under the Open Database License, and the full eICU database to credentialed PhysioNet users.
The MedQA and Medbullets questions are available from their original sources, and the code rebuilds the survey and intensive care records and the board questions from these sources. The patient questions, pathology reports, and model responses are available from the corresponding author on reasonable request.
Our code, prompt templates and aggregate results are available at https://github.com/deanariel-md/decision-model-offload and archived at https://doi.org/10.5281/zenodo.23070012.

## Statistical analysis

Systems were compared on the same items with 2,000 paired bootstrap resamples, stratified by question type, stage level, or, for cancer staging, report type. Non-inferiority of Jev, and of each pairing, against each of the ten LLMs was tested one-sided at α = 0.025 with a margin of 5 percentage points and Holm’s adjustment across the ten^35^ (for the pairings, Bonferroni’s, which is stricter): a comparison was non-inferior when the adjusted lower bound of the difference lay above −5 points, worse when the upper bound lay below it, and inconclusive otherwise. Risk of death in NHANES used log-loss with the margin above and simultaneous 95% intervals across the ten LLMs (maximum-statistic method); comparisons in intensive care, which has no such margin, were descriptive. Repeated calls on identical input are summarised in Supplementary Table 11. Analyses used Python 3.12 with pandas 2.3.3, NumPy 2.4.4, SciPy 1.17.1, scikit-learn 1.7.2, LightGBM 4.6.0, and XGBoost 3.2.0.

## Ethics

Only de-identified data were used: the NHANES public-use files, the open-access eICU demo, and the credentialed eICU database under its data use agreement^10^. The patient questions and pathology reports describe no real patients. No further ethical review was required.

## Data availability

NHANES and the Linked Mortality Files are available from the National Center for Health Statistics, the eICU demo from PhysioNet under the Open Database License, and the full eICU database to credentialed PhysioNet users. The MedQA and Medbullets questions are available from their original sources, and the code rebuilds the survey and intensive care records and the board questions from these sources. The patient questions, pathology reports, and model responses are available from the corresponding author on reasonable request.

## Code availability

Code to build the prompts, query the systems, and analyse the replies, with the prompt templates and aggregate results, is available at https://github.com/deanariel-md/decision-model-offload and archived at Zenodo (https://doi.org/10.5281/zenodo.23070012).

## Declarations

### Author contributions

D.A. conceived the study, built the datasets and analyses, and wrote the manuscript. D.A., G.L., R.B., N.D., and S.C. planned and designed the study. S.E. validated the patient questions. N.D. and S.C. supervised the study. All authors reviewed and approved the manuscript.

