## Supplementary Appendix for "Decision models can offload structured clinical tasks from large language models"

#### Contents


| | |
|---|---|
| Supplementary Fig. 1 | Confidence thresholds: accuracy and cost of the pairing |
| Supplementary Table 1 | Ten-year risk of death: estimates of every system |
| Supplementary Table 2 | Choosing Wisely patient questions and board questions, every system |
| Supplementary Table 3 | Cancer and kidney-injury staging, every system |
| Supplementary Table 4 | Cost and response time |
| Supplementary Table 5 | Cancer staging: every form of question, every system (240 reports) |
| Supplementary Table 6 | Confidence and correctness |
| Supplementary Table 7 | Jev with an LLM |
| Supplementary Table 8 | Systems tested |
| Supplementary Table 9 | Ten-year risk: cohort and participants |
| Supplementary Table 10 | Ten-year risk: response to one-field edits |
| Supplementary Table 11 | Repeatability and prior exposure |
| Supplementary Table 12 | Risk of death: the named categories of the recommended form |
| Supplementary Table 13 | Clinical decisions: sensitivity, specificity and AUROC |
| Supplementary Note 1 | Prompts and reading of replies |
| Supplementary Note 2 | Recommended form for cancer staging: the questions and options as sent |
| Supplementary Note 3 | Outcome reference |
| Supplementary Note 4 | Cancer staging: how the reports were built |

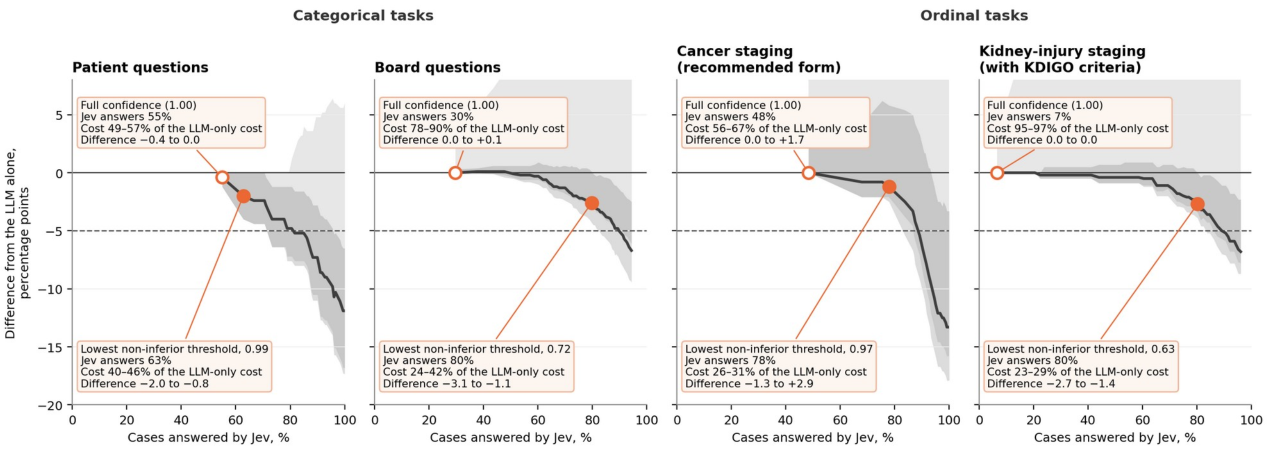


### Supplementary Fig. 1. Confidence thresholds: accuracy and cost of the pairing

Jev answers the items whose confidence reaches the threshold and passes the rest to the LLM; lowering the threshold moves right (Jev answers more, and the cost falls). Line, median difference from the LLM alone across the five flagship LLMs; dark grey band, from the lowest to the highest bound of their 95% bootstrap intervals; light grey band, the same for the five smaller LLMs; dashed, the 5-point non-inferiority margin. Open dot, full confidence (the threshold of the main analysis); filled dot, the lowest threshold at which the pairing with each of the ten LLMs was non-inferior (one-sided, Bonferroni over the ten LLMs; thresholds examined descriptively). Box at each dot: Jev’s share of the items and, across the five flagship LLMs, the range of cost and of the difference. Balanced accuracy for patient questions, exact accuracy otherwise. Cancer staging: the 240 pathology reports, Jev in the recommended form, the LLMs with stage names. Values at each threshold: Supplementary Table 7c.

### Supplementary Table 1. Ten-year risk of death: estimates of every system

#### a. Every system, standard LLM-style inputs

| Predictor | Log-loss (95% CI) | Recalibrated log-loss | Brier score (95% CI) | AUROC | Calibration slope | Calibration intercept | Mean predicted risk, % |
|---|---|---|---|---|---|---|---|
| Jev 1.13 | 0.379 (0.372–0.386) | 0.338 | 0.117 (0.114–0.120) | 0.822 | 1.14 | −0.74 | 26.3 |
| GPT-5.6 Sol | 0.329 (0.321–0.337) | 0.325 | 0.101 (0.098–0.104) | 0.839 | 1.01 | −0.28 | 18.8 |
| Claude Opus 5.5 | 0.327 (0.318–0.335) | 0.319 | 0.099 (0.097–0.102) | 0.846 | 1.10 | −0.28 | 19.8 |
| Gemini 3.8 Flash | 0.325 (0.317–0.334) | 0.322 | 0.099 (0.096–0.103) | 0.842 | 1.01 | −0.26 | 18.5 |
| Muse Spark 1.1 | 0.362 (0.354–0.369) | 0.327 | 0.112 (0.110–0.115) | 0.837 | 1.03 | −0.78 | 25.2 |
| GLM-5.3 | 0.330 (0.322–0.339) | 0.327 | 0.101 (0.098–0.104) | 0.837 | 1.09 | −0.17 | 18.6 |
| Reference, spline logistic | 0.311 (0.301–0.321) | 0.311 | 0.094 (0.091–0.098) | 0.854 | 0.93 | −0.08 | 15.7 |
| Reference, LightGBM | 0.313 (0.303–0.324) | 0.313 | 0.095 (0.092–0.098) | 0.851 | 0.91 | −0.06 | 15.3 |
| PhenoAge[1] | 0.364 (0.351–0.377) | 0.351 | 0.109 (0.105–0.114) | 0.812 | 0.77 | 0.01 | 12.0 |
| Age and sex only | 0.371 (0.362–0.382) | 0.372 | 0.114 (0.111–0.118) | 0.769 | 0.99 | −0.02 | 15.8 |

12,697 records answered by every system (2,002 deaths, 15.8%): the evaluation set and the fitting-set records with all nine biomarkers, the latter scored against ten-fold out-of-fold reference predictions. Probabilities clipped to [0.005, 0.995]. Recalibration: cross-fitted logistic recalibration.

Log-loss difference, Jev − LLM (simultaneous 95% CI over the ten LLMs; non-inferiority margin 0.0060):

| LLM | Difference | Non-inferiority |
|---|---|---|
| GPT-5.6 Sol | +0.050 (+0.045 to +0.055) | worse |
| Claude Opus 5.5 | +0.052 (+0.048 to +0.057) | worse |
| Gemini 3.8 Flash | +0.053 (+0.048 to +0.059) | worse |
| Muse Spark 1.1 | +0.017 (+0.014 to +0.021) | worse |
| GLM-5.3 | +0.048 (+0.043 to +0.054) | worse |
| GPT-5.6 Luna | +0.041 (+0.037 to +0.045) | worse |
| Claude Sonnet 5 | +0.025 (+0.022 to +0.029) | worse |
| Gemini 3.5 Flash-Lite | +0.041 (+0.036 to +0.045) | worse |
| MedGemma 27B | +0.009 (−0.001 to +0.019) | inconclusive |
| Gemma 3 27B | +0.014 (+0.005 to +0.023) | inconclusive |

**b. Jev in the recommended form**

Baseline prediction, 12,697 participants, 2,002 deaths:

| System | Log-loss (95% CI) | Recalibrated log-loss | Brier score | AUROC | Calibration slope | Mean estimate, % |
|---|---|---|---|---|---|---|
| Jev 1.13, recommended form | 0.362 (0.355–0.369) | 0.339 | 0.110 | 0.820 | 1.23 | 23.2 |
| Jev 1.13, standard LLM-style inputs | 0.379 (0.372–0.386) | 0.338 | 0.117 | 0.822 | 1.14 | 26.3 |
| GPT-5.6 Sol | 0.329 (0.321–0.337) | 0.325 | 0.101 | 0.839 | 1.01 | 18.8 |
| Claude Opus 5.5 | 0.327 (0.318–0.335) | 0.319 | 0.099 | 0.846 | 1.10 | 19.8 |
| Gemini 3.8 Flash | 0.325 (0.317–0.334) | 0.322 | 0.099 | 0.842 | 1.01 | 18.5 |
| Muse Spark 1.1 | 0.362 (0.354–0.369) | 0.327 | 0.112 | 0.837 | 1.03 | 25.2 |
| GLM-5.3 | 0.330 (0.322–0.339) | 0.327 | 0.101 | 0.837 | 1.09 | 18.6 |
| GPT-5.6 Luna | 0.338 (0.330–0.346) | 0.329 | 0.103 | 0.834 | 1.16 | 20.2 |
| Claude Sonnet 5 | 0.353 (0.346–0.361) | 0.332 | 0.108 | 0.830 | 1.18 | 23.0 |
| Gemini 3.5 Flash-Lite | 0.338 (0.330–0.347) | 0.331 | 0.103 | 0.832 | 1.17 | 19.7 |
| MedGemma 27B | 0.370 (0.358–0.381) | 0.367 | 0.113 | 0.783 | 0.80 | 14.4 |
| Gemma 3 27B | 0.365 (0.355–0.374) | 0.365 | 0.113 | 0.787 | 1.07 | 15.6 |
| Reference, spline logistic | 0.311 (0.301–0.321) | 0.311 | 0.094 | 0.854 | 0.93 | 15.7 |
| Reference, LightGBM | 0.313 (0.303–0.324) | 0.313 | 0.095 | 0.851 | 0.91 | 15.3 |
| Age and sex only | 0.371 (0.362–0.382) | 0.372 | 0.114 | 0.769 | 0.99 | 15.8 |
| PhenoAge | 0.364 (0.351–0.377) | 0.351 | 0.109 | 0.812 | 0.77 | 12.0 |

Jev in the recommended form minus each system, log-loss (simultaneous 95% CI; for the LLMs, over the ten):

| Compared with | Log-loss difference | Non-inferiority |
|---|---|---|
| Jev 1.13, standard LLM-style inputs | −0.017 (−0.019 to −0.015) | — |
| GPT-5.6 Sol | +0.033 (+0.029 to +0.037) | worse |
| Claude Opus 5.5 | +0.035 (+0.031 to +0.039) | worse |
| Gemini 3.8 Flash | +0.036 (+0.032 to +0.041) | worse |
| Muse Spark 1.1 | +0.0002 (−0.004 to +0.005) | non-inferior |
| GLM-5.3 | +0.031 (+0.027 to +0.036) | worse |
| GPT-5.6 Luna | +0.024 (+0.020 to +0.027) | worse |
| Claude Sonnet 5 | +0.008 (+0.005 to +0.012) | inconclusive |
| Gemini 3.5 Flash-Lite | +0.024 (+0.020 to +0.027) | worse |
| MedGemma 27B | −0.008 (−0.017 to +0.001) | non-inferior |
| Gemma 3 27B | −0.003 (−0.010 to +0.005) | non-inferior |
| Age and sex only | −0.010 (−0.016 to −0.004) | non-inferior |
| Reference, spline logistic | +0.050 (+0.043 to +0.058) | worse |

Lower log-loss is better. The LLMs' answers, the references, the records and the analysis are those of part a; only Jev was called again, with every number replaced by its named category (Supplementary Table 12). Non-inferiority margin 0.0060 log-loss (10% of the gap between the age-and-sex model and the reference). The smaller LLMs were scored on the same records with the same code. Simultaneous intervals for the LLMs: maximum-statistic over the ten, 2,000 resamples of the 12,697 records. Jev answered 12,910 of 12,910 requests usably, with 825 input tokens per request on average, at US$0.035 per 1,000 at list price.

#### c. Discrimination and calibration in the large

| System | AUROC (95% CI) | Jev (standard LLM-style inputs) − system, AUROC (95% CI) | Mean predicted risk, % | Observed ÷ predicted | Calibration slope |
|---|---|---|---|---|---|
| Jev, standard LLM-style inputs | 0.822 (0.813–0.832) | — | 26.3 | 0.60 | 1.14 |
| Jev, recommended form | 0.820 (0.811–0.831) | +0.002 (−0.001 to +0.005) | 23.2 | 0.68 | 1.23 |
| GPT-5.6 Sol | 0.839 (0.829–0.848) | −0.016 (−0.020 to −0.012) | 18.8 | 0.84 | 1.01 |
| Claude Opus 5.5 | 0.846 (0.837–0.855) | −0.023 (−0.027 to −0.019) | 19.8 | 0.79 | 1.10 |
| Gemini 3.8 Flash | 0.842 (0.833–0.851) | −0.020 (−0.024 to −0.016) | 18.5 | 0.85 | 1.01 |
| Muse Spark 1.1 | 0.837 (0.828–0.847) | −0.015 (−0.018 to −0.011) | 25.2 | 0.63 | 1.03 |
| GLM-5.3 | 0.837 (0.828–0.846) | −0.015 (−0.019 to −0.011) | 18.6 | 0.85 | 1.09 |
| Reference model (spline logistic regression) | 0.854 (0.845–0.863) | −0.032 (−0.038 to −0.026) | 15.7 | 1.01 | 0.93 |
| Age and sex only | 0.769 (0.758–0.780) | +0.053 (+0.046 to +0.061) | 15.8 | 1.00 | 0.99 |
| GPT-5.6 Luna | 0.834 (0.825–0.844) | — | — | — | — |
| Claude Sonnet 5 | 0.830 (0.821–0.840) | — | — | — | — |
| Gemini 3.5 Flash-Lite | 0.832 (0.823–0.842) | — | — | — | — |
| MedGemma 27B | 0.783 (0.772–0.794) | — | — | — | — |
| Gemma 3 27B | 0.787 (0.777–0.798) | — | — | — | — |

The 12,697 baseline records every system answered; 2,002 (15.8%) died within ten years. Intervals: 2,000 bootstrap resamples of records, paired for the differences. References scored out of fold. Smaller LLMs: AUROC with the same bootstrap; their other measures are in parts a and b.

#### d. Replication: death in hospital, eICU demo

| Predictor | Log-loss (95% CI) | Brier score (95% CI) | AUROC (95% CI) | Calibration slope (95% CI) | Mean predicted risk, % | Observed to expected (95% CI) |
|---|---|---|---|---|---|---|
| Jev 1.13 | 0.375 (0.359–0.391) | 0.112 (0.106–0.119) | 0.836 (0.795–0.872) | 1.71 (1.41–2.06) | 28.9 | 0.26 (0.22–0.31) |
| GPT-5.6 Sol | 0.210 (0.190–0.232) | 0.058 (0.051–0.066) | 0.874 (0.838–0.904) | 1.28 (1.07–1.54) | 13.2 | 0.57 (0.48–0.67) |
| Claude Opus 5.5 | 0.213 (0.194–0.234) | 0.058 (0.051–0.066) | 0.880 (0.845–0.909) | 1.44 (1.23–1.70) | 13.7 | 0.55 (0.46–0.65) |
| Gemini 3.8 Flash | 0.251 (0.233–0.271) | 0.071 (0.064–0.079) | 0.864 (0.828–0.895) | 1.22 (1.03–1.43) | 18.0 | 0.42 (0.35–0.50) |
| Muse Spark 1.1 | 0.264 (0.245–0.285) | 0.076 (0.068–0.084) | 0.864 (0.828–0.896) | 1.17 (1.00–1.39) | 19.4 | 0.39 (0.32–0.46) |
| GLM-5.3 | 0.297 (0.279–0.315) | 0.084 (0.077–0.091) | 0.845 (0.806–0.881) | 1.34 (1.14–1.58) | 22.0 | 0.34 (0.28–0.41) |
| GPT-5.6 Luna | 0.267 (0.248–0.287) | 0.077 (0.069–0.084) | 0.856 (0.816–0.892) | 1.17 (0.97–1.42) | 19.1 | 0.39 (0.33–0.47) |
| Claude Sonnet 5 | 0.257 (0.238–0.276) | 0.072 (0.065–0.079) | 0.860 (0.822–0.894) | 1.33 (1.12–1.59) | 18.4 | 0.41 (0.34–0.48) |
| Gemini 3.5 Flash-Lite | 0.290 (0.274–0.307) | 0.082 (0.075–0.089) | 0.854 (0.816–0.888) | 1.64 (1.34–2.01) | 21.7 | 0.35 (0.29–0.41) |
| MedGemma 27B | 0.628 (0.579–0.679) | 0.202 (0.187–0.217) | 0.795 (0.751–0.832) | 0.49 (0.40–0.59) | 35.9 | 0.21 (0.17–0.25) |
| Gemma 3 27B | 0.558 (0.514–0.602) | 0.178 (0.164–0.192) | 0.808 (0.767–0.844) | 0.52 (0.43–0.63) | 33.3 | 0.23 (0.19–0.27) |
| APACHE IVa | 0.198 (0.173–0.224) | 0.055 (0.046–0.064) | 0.864 (0.828–0.893) | 1.07 (0.92–1.27) | 9.8 | 0.77 (0.65–0.91) |
| Reference, spline logistic | 0.183 (0.155–0.213) | 0.052 (0.043–0.062) | 0.888 (0.857–0.917) | 1.15 (0.98–1.37) | 6.8 | 1.11 (0.94–1.31) |
| Reference, LightGBM | 0.178 (0.151–0.208) | 0.050 (0.042–0.060) | 0.892 (0.859–0.921) | 1.22 (1.04–1.45) | 6.7 | 1.13 (0.95–1.33) |
| Reference, XGBoost | 0.179 (0.152–0.209) | 0.051 (0.042–0.060) | 0.894 (0.860–0.922) | 1.28 (1.09–1.51) | 6.6 | 1.14 (0.96–1.33) |

1,246 stays answered by every predictor (94 deaths, 7.5%), of 1,297 evaluation stays. References fitted on the full eICU database without any demo patient.

**e. Replication: death in hospital, Jev in the recommended form**

| Predictor | Log-loss (95% CI) | Brier score (95% CI) | AUROC (95% CI) | Calibration slope (95% CI) | Mean predicted risk, % | Observed to expected (95% CI) |
|---|---|---|---|---|---|---|
| Jev 1.13, standard LLM-style inputs | 0.373 (0.358–0.388) | 0.111 (0.105–0.118) | 0.837 (0.797–0.875) | 1.72 (1.42–2.08) | 28.7 | 0.25 (0.21–0.30) |
| Jev 1.13, recommended form | 0.311 (0.296–0.327) | 0.087 (0.081–0.093) | 0.818 (0.779–0.855) | 1.62 (1.34–1.96) | 22.6 | 0.32 (0.26–0.38) |

1,297 evaluation stays, 95 deaths (7.3%). AUROC, recommended form minus standard LLM-style inputs: −0.019 (−0.043 to +0.006) (paired DeLong, P = 0.13). Categories: Supplementary Table 12.

### Supplementary Table 2. Choosing Wisely patient questions and board questions, every system

#### a. Patient questions: is the test or treatment needed? (180)

| System | Balanced accuracy, % (95% CI) | Low-value questions correct, % | Indicated questions correct, % | Overuse, % | False restraint, % | Other answer, % | Unparsable answers | Jev − system, points (95% CI) | Non-inferiority of Jev |
|---|---|---|---|---|---|---|---|---|---|
| Jev 1.13 | 87.7 (82.7–92.1) | 80.8 | 94.5 | 19.2 | 5.5 | 0.0 | 0 | | |
| GPT-5.6 Sol | 100.0 | 100.0 | 100.0 | 0.0 | 0.0 | 0.0 | 0 | −12.3 (−17.3 to −7.9) | worse |
| Claude Opus 5.5 | 100.0 | 100.0 | 100.0 | 0.0 | 0.0 | 0.0 | 0 | −12.3 (−17.3 to −7.9) | worse |
| Gemini 3.8 Flash | 99.6 (98.8–100.0) | 99.2 | 100.0 | 0.8 | 0.0 | 0.0 | 0 | −11.9 (−16.9 to −7.6) | worse |
| Muse Spark 1.1 | 99.1 (97.3–100.0) | 100.0 | 98.2 | 0.0 | 1.8 | 0.0 | 0 | −11.4 (−16.7 to −6.6) | inconclusive |
| GLM-5.3 | 98.4 (96.8–99.6) | 96.8 | 100.0 | 3.2 | 0.0 | 0.0 | 0 | −10.7 (−15.6 to −6.5) | worse |
| GPT-5.6 Luna | 99.6 (98.8–100.0) | 99.2 | 100.0 | 0.8 | 0.0 | 0.0 | 0 | −11.9 (−16.8 to −7.6) | worse |
| Claude Sonnet 5 | 97.5 (94.9–99.6) | 96.8 | 98.2 | 3.2 | 1.8 | 0.0 | 0 | −9.8 (−14.7 to −5.1) | inconclusive |
| Gemini 3.5 Flash-Lite | 95.5 (92.2–98.0) | 92.8 | 98.2 | 7.2 | 1.8 | 0.0 | 0 | −7.8 (−12.1 to −4.0) | inconclusive |
| MedGemma 27B | 86.1 (80.8–90.4) | 77.6 | 94.5 | 22.4 | 5.5 | 0.0 | 0 | +1.6 (−2.2 to +5.3) | non-inferior |
| Gemma 3 27B | 85.0 (80.1–89.2) | 73.6 | 96.4 | 26.4 | 3.6 | 0.0 | 0 | +2.7 (−0.9 to +6.0) | non-inferior |
| Style-only baseline | 69.3 (62.1–76.2) | 64.0 | 74.5 | | | | | | |

180 patient questions. Overuse: the test or treatment chosen in a low-value question; false restraint: holding off chosen in an indicated question; other answer: one of the two actions inappropriate in either kind of question. Differences from paired bootstrap resamples stratified by question type; non-inferiority with Holm’s adjustment across the ten LLMs, margin 5 points. The questions of one recommendation share an answer set, so the 14 recommendations were also resampled whole (2,000 resamples). With whole recommendations resampled, Jev was inconclusive against GPT-5.6 Sol, Claude Opus 5.5, Gemini 3.8 Flash, Muse Spark 1.1, GLM-5.3, GPT-5.6 Luna, Claude Sonnet 5 and Gemini 3.5 Flash-Lite (upper bounds −2.9 to −1.2 points, lower bounds −23.6 to −15.1) and non-inferior to MedGemma 27B and Gemma 3 27B.

Balanced accuracy, % (95% CI), on the 121 questions the style-only baseline classified correctly and the 59 it did not:

| System | Style points to the answer | Style points away from the answer |
|---|---|---|
| Jev 1.13 | 85.7 (79.5–91.2) | 90.9 (81.5–97.8) |
| GPT-5.6 Sol | 100.0 | 100.0 |
| Claude Opus 5.5 | 100.0 | 100.0 |
| Gemini 3.8 Flash | 100.0 | 98.9 (96.7–100.0) |
| Muse Spark 1.1 | 98.8 (96.3–100.0) | 100.0 |
| GLM-5.3 | 98.1 (95.6–100.0) | 98.9 (96.7–100.0) |
| GPT-5.6 Luna | 99.4 (98.1–100.0) | 100.0 |
| Claude Sonnet 5 | 98.1 (95.6–100.0) | 95.3 (87.1–100.0) |
| Gemini 3.5 Flash-Lite | 93.8 (89.4–97.5) | 98.9 (96.7–100.0) |
| MedGemma 27B | 84.4 (78.8–90.0) | 88.7 (79.3–95.6) |
| Gemma 3 27B | 83.8 (78.2–88.8) | 86.4 (77.1–94.4) |

**b. Board questions**

| System | All, % (95% CI) | MedQA, % | Medbullets, % | Management questions, % | Other option count, % | Unparsable answers | Jev − system, points (95% CI) | Non-inferiority of Jev |
|---|---|---|---|---|---|---|---|---|
| Jev 1.13 | 85.6 (83.9–87.3) | 88.1 (86.3–89.9) | 75.2 (70.1–79.9) | 81.3 (77.2–85.2) | 83.5 (81.7–85.4) | 0 | | |
| GPT-5.6 Sol | 94.7 (93.6–95.8) | 96.0 (94.9–97.0) | 89.3 (85.6–92.6) | 91.9 (88.9–94.4) | 94.0 (92.8–95.3) | 0 | −9.1 (−10.9 to −7.5) | worse |
| Claude Opus 5.5 | 95.9 (94.9–96.9) | 97.2 (96.3–98.0) | 90.3 (86.9–93.6) | 93.9 (91.4–96.1) | 95.0 (93.9–96.1) | 3 | −10.2 (−12.0 to −8.5) | worse |
| Gemini 3.8 Flash | 95.7 (94.7–96.7) | 96.9 (95.9–97.8) | 90.6 (87.2–93.6) | 92.2 (89.4–94.7) | 95.5 (94.5–96.6) | 0 | −10.1 (−11.8 to −8.4) | worse |
| Muse Spark 1.1 | 92.8 (91.5–94.0) | 94.1 (92.7–95.4) | 87.2 (83.6–90.6) | 89.4 (86.1–92.5) | 92.3 (91.0–93.5) | 7 | −7.2 (−9.0 to −5.5) | inconclusive |
| GLM-5.3 | 91.2 (89.8–92.6) | 92.8 (91.3–94.2) | 84.6 (80.2–88.6) | 88.9 (85.5–91.9) | 90.7 (89.2–92.2) | 44 | −5.6 (−7.3 to −3.9) | inconclusive |
| GPT-5.6 Luna | 91.9 (90.6–93.2) | 94.0 (92.7–95.3) | 82.9 (78.5–86.9) | 88.9 (85.5–91.9) | 92.0 (90.6–93.3) | 0 | −6.3 (−8.0 to −4.8) | inconclusive |
| Claude Sonnet 5 | 92.2 (90.9–93.5) | 93.9 (92.6–95.2) | 85.2 (80.9–88.9) | 90.0 (86.6–92.8) | 92.0 (90.6–93.4) | 2 | −6.6 (−8.3 to −5.0) | inconclusive |
| Gemini 3.5 Flash-Lite | 86.1 (84.5–87.8) | 88.3 (86.5–90.0) | 76.8 (72.1–81.5) | 82.7 (78.6–86.6) | 84.7 (82.9–86.6) | 0 | −0.5 (−2.2 to +1.2) | non-inferior |
| MedGemma 27B | 68.2 (65.8–70.5) | 71.4 (68.9–73.9) | 54.4 (48.7–59.7) | 65.7 (60.7–70.5) | 66.5 (64.2–68.8) | 0 | +17.4 (+15.3 to +19.5) | non-inferior |
| Gemma 3 27B | 63.4 (61.1–65.8) | 67.3 (64.8–69.8) | 46.6 (40.9–52.3) | 59.3 (54.0–64.3) | 60.5 (58.2–63.0) | 0 | +22.2 (+20.0 to +24.5) | non-inferior |

1,571 questions (MedQA 1,273, Medbullets 298). Jev minus each system: paired bootstrap stratified by source, Holm's adjustment across the ten LLMs, margin 5 points; management questions: the 359 whose final sentence asked what to do next, the most appropriate management or the best treatment. Other option count: every question asked again with five options for MedQA and four for Medbullets. Jev gave a probability of 1.00 to 465 questions (29.6%) and was correct on 99.6% of them, against 79.7% on the rest.

**c. The 14 Choosing Wisely recommendations and their answer sets**

| | Recommendation | A. Test or treatment | B. Holding off | C (wrong in both) | D (wrong in both) | Low-value | Indicated |
|---|---|---|---|---|---|---|---|
| 1 | Imaging for low back pain | Arrange imaging of the lumbar spine (MRI or CT) now. | No imaging for now: stay active, use simple pain relief, and review if the pain or symptoms change. | Start a short course of oral steroids and review in two weeks. | Start an opioid painkiller and review in two weeks. | 10 | 5 |
| 2 | Antibiotics for upper respiratory infection | Start antibiotic treatment now (intravenous if severe). | No antibiotic: symptom relief and review if the illness does not follow the expected course. | Order a chest X-ray. | Prescribe a five-day course of oral steroids. | 9 | 2 |
| 3 | PSA screening at 70 or older | Order a PSA blood test. | No PSA test: routine screening is not recommended at this age; review if new or worsening symptoms appear. | Start tamsulosin and review in three months. | Order a urine culture and prescribe antibiotics. | 4 | 2 |
| 4 | Thyroid ultrasound for an incidental subcentimetre nodule | Arrange a thyroid ultrasound. | No thyroid imaging for now; review if a lump or new neck symptoms appear. | Start levothyroxine. | Arrange a radioiodine (nuclear) thyroid scan. | 10 | 3 |
| 5 | Vitamin D testing | Order a serum vitamin D (25-hydroxyvitamin D) level. | No vitamin D test: a balanced diet and usual activity; review if symptoms develop. | Order thyroid function tests. | Order a serum vitamin B12 level. | 8 | 5 |
| 6 | Head CT after fainting | Arrange a CT scan of the head now. | No head scan: an ECG, fluids and avoiding known triggers; review if fainting recurs or new symptoms appear. | Arrange a tilt-table test. | Start an anti-seizure medicine. | 10 | 5 |
| 7 | Resting ECG | Arrange a resting 12-lead ECG. | No ECG: routine care and review if new symptoms appear. | Arrange a carotid ultrasound scan. | Order a D-dimer blood test. | 7 | 5 |
| 8 | Preoperative testing | Order preoperative tests (blood tests, chest X-ray and/or ECG) before surgery. | No routine preoperative tests; proceed as planned after the standard history and examination. | Order a routine CT scan of the lungs before surgery. | Start a beta-blocker the day before surgery. | 10 | 5 |
| 9 | Bone density (DEXA) under 65 without risk factors | Arrange a DEXA bone density scan. | No bone density scan for now: weight-bearing exercise, enough calcium and vitamin D, and screen at 65. | Arrange a heel ultrasound bone test instead of DEXA. | Start hormone replacement therapy to prevent osteoporosis. | 9 | 2 |
| 10 | Brain imaging for headache | Arrange brain imaging (CT or MRI) now. | No brain scan: treatment matched to the headache type, a headache diary, and review if the headache pattern changes. | Arrange an EEG. | Prescribe an opioid painkiller for the headache. | 9 | 5 |
| 11 | Opioids for low back pain | Prescribe an opioid painkiller. | No opioid: heat, staying active and non-opioid pain relief suited to the person; review if not improving. | Prescribe a benzodiazepine for muscle spasm. | Order an MRI and give no pain relief until it is done. | 10 | 4 |
| 12 | Benzodiazepines at 65 or older | Prescribe (or continue) a benzodiazepine. | No benzodiazepine: non-drug measures and treat the underlying cause; review. | Prescribe zolpidem instead. | Prescribe diphenhydramine (an antihistamine sleep aid). | 10 | 3 |
| 13 | Antibiotics for acute sinusitis under 10 days | Start antibiotic treatment now (for example amoxicillin-clavulanate; intravenous if severe). | No antibiotic: saline irrigation and pain relief; review if the illness does not follow the expected course. | Prescribe an oral steroid on its own. | Prescribe an oral antihistamine. | 10 | 5 |
| 14 | CT pulmonary angiography | Arrange a CT pulmonary angiogram now. | No CT scan: pulmonary embolism is unlikely on these findings; treat the likely cause and review. | Order a D-dimer blood test. | Prescribe an antibiotic for a possible chest infection. | 9 | 4 |
| | **Total** | | | | | 125 | 55 |

One answer set per recommendation, identical for every system; the order of the four answers was shuffled once per question. Key: B for a low-value question, A for an indicated one; C and D are wrong in both. Low-value and indicated: number of questions.

### Supplementary Table 3. Cancer and kidney-injury staging, every system

#### a. Stage levels (AJCC 8th edition), 240 reports

| Level | T, N and M | Reports |
|---|---|---|
| 0 | Tis N0 M0 | 15 |
| I | T1–T2 N0 M0 | 21 |
| IIA | T3 N0 M0 | 26 |
| IIB | T4a N0 M0 | 38 |
| IIC | T4b N0 M0 | 16 |
| IIIA | T1–T2 N1a–N1c M0; T1 N2a M0 | 12 |
| IIIB | T3–T4a N1a–N1c M0; T2–T3 N2a M0; T1–T2 N2b M0 | 25 |
| IIIC | T4a N2a M0; T3–T4a N2b M0; T4b N1a–N2b M0 | 26 |
| IVA–IVB | Any T, any N, M1a–M1b | 53 (45 IVA, 8 IVB) |
| IVC | Any T, any N, M1c | 8 |

The 240 reports: 120 ordinary and 120 built to mislead (100 with one misleading feature and 20 with a metastatic node at an unusual site; Supplementary Note 4). N1c: tumour deposits with no involved regional lymph node. M1a: metastasis to one site or organ, without peritoneal metastasis; M1b: to two or more sites or organs, without peritoneal metastasis; M1c: to the peritoneal surface, alone or with other sites or organs.

#### b. Cancer staging, every system, same text for all systems

| System | Stage names only, % (95% CI) | With medical context, % (95% CI) | Quadratic weighted kappa | Ranked probability score | Unparsable answers (stage names / medical context) | Jev − system, stage names, points (95% CI) | Non-inferiority of Jev |
|---|---|---|---|---|---|---|---|
| Jev 1.13 | 45.4 (39.2–51.7) | 75.8 (70.0–80.8) | 0.941 | 0.054 | 0 / 0 | | |
| GPT-5.6 Sol | 99.2 (97.0–99.8) | 99.6 (97.7–99.9) | 0.999 | 0.001 | 0 / 0 | −53.8 (−60.0 to −47.5) | worse |
| Claude Opus 5.5 | 97.1 (94.1–98.6) | 100.0 | 0.998 | 0.004 | 0 / 0 | −51.7 (−58.3 to −45.4) | worse |
| Gemini 3.8 Flash | 100.0 | 100.0 | 1.000 | 0.000 | 0 / 0 | −54.6 (−60.8 to −48.3) | worse |
| Muse Spark 1.1 | 99.6 (97.7–99.9) | 100.0 | 1.000 | 0.001 | 0 / 0 | −54.2 (−60.4 to −47.9) | worse |
| GLM-5.3 | 94.2 (90.4–96.5) | 97.5 (94.7–98.8) | 0.999 | 0.002 | 11 / 6 | −48.8 (−55.4 to −41.7) | worse |
| GPT-5.6 Luna | 97.1 (94.1–98.6) | 96.7 (93.6–98.3) | 0.998 | 0.003 | 0 / 0 | −51.7 (−58.3 to −44.6) | worse |
| Claude Sonnet 5 | 97.5 (94.7–98.8) | 95.8 (92.5–97.7) | 0.998 | 0.004 | 0 / 0 | −52.1 (−59.2 to −45.4) | worse |
| Gemini 3.5 Flash-Lite | 59.2 (52.9–65.2) | 73.8 (67.8–78.9) | 0.959 | 0.049 | 0 / 0 | −13.8 (−21.7 to −5.8) | inconclusive |
| MedGemma 27B | 30.0 (24.6–36.1) | 39.2 (33.2–45.5) | 0.677 | 0.162 | 0 / 0 | +15.4 (+7.9 to +22.9) | non-inferior |
| Gemma 3 27B | 26.7 (21.5–32.6) | 25.8 (20.7–31.7) | 0.685 | 0.127 | 0 / 0 | +18.8 (+11.2 to +25.8) | non-inferior |

Exact accuracy on the 240 reports; Wilson 95% intervals. Kappa and ranked probability score refer to the stage-names version (ranked probability score: lower is better). Jev minus each system: 2,000 paired bootstrap resamples stratified by report type; non-inferiority margin 5 points, one-sided α = 0.025 with Holm's adjustment across the ten LLMs. In neither version did three or more of the five flagship LLMs agree on a stage other than the key for any report. Jev's highest probability for any level was 0.83 with the stage names only. Every system in every form of the question: Supplementary Table 5.

#### c. Cancer staging, Jev in the recommended form

| Version | Stage group exact, % (95% CI) | T category, % | Involved nodes, % | Tumour deposits, % | Distant metastasis, % | Stage III answered 0–II | Unparsable answers | Input tokens per answer | List cost per 1,000 answers, US$ |
|---|---|---|---|---|---|---|---|---|---|
| Recommended form | 85.8 (80.9–89.7) | 100.0 | 100.0 | 100.0 | 85.8 | 0 of 63 | 0 | 2,776 | 0.117 |

240 reports. Four Choice questions in one request (T category; involved regional nodes in bands; tumour deposits; distant metastasis), each option with its AJCC 8th edition definition, what it excludes and examples that appear in no report; the stage group computed in code from the four answers with the table that built the key (Supplementary Note 2). Response time, 100 reports sent one at a time: median 0.79 s (90th percentile 0.89 s).

#### d. Kidney-injury staging, every system

| System | Stage names only, % (95% CI) | With KDIGO criteria, % (95% CI) | Balanced accuracy, stage names, % | Balanced accuracy, KDIGO criteria, % | Stage 2 or 3 answered 2 or 3, stage names, % | Stage 2 or 3 answered 2 or 3, KDIGO criteria, % | Unparsable answers, stage names / KDIGO criteria | Jev − system, names, points (95% CI) | Non-inferiority of Jev |
|---|---|---|---|---|---|---|---|---|---|
| Jev 1.13 | 54.4 (50.6–58.1) | 91.8 (89.3–94.1) | 60.5 | 91.4 | 94.0 | 95.2 | 0 / 0 | | |
| GPT-5.6 Sol | 95.4 (93.6–97.0) | 100.0 | 97.0 | 100.0 | 98.8 | 100.0 | 0 / 0 | −41.0 (−45.3 to −36.9) | worse |
| Claude Opus 5.5 | 99.5 (98.8–100.0) | 99.8 (99.5–100.0) | 98.9 | 99.2 | 97.6 | 98.8 | 0 / 0 | −45.1 (−48.8 to −41.5) | worse |
| Gemini 3.8 Flash | 96.1 (94.3–97.5) | 100.0 | 97.1 | 100.0 | 98.8 | 100.0 | 0 / 0 | −41.7 (−45.8 to −38.0) | worse |
| Muse Spark 1.1 | 98.2 (97.0–99.1) | 100.0 | 97.5 | 100.0 | 96.4 | 100.0 | 0 / 0 | −43.9 (−47.6 to −40.1) | worse |
| GLM-5.3 | 90.9 (88.4–93.0) | 97.9 (96.4–98.9) | 89.7 | 97.7 | 88.0 | 97.6 | 47 / 12 | −36.5 (−40.8 to −32.4) | worse |
| GPT-5.6 Luna | 91.8 (89.7–93.9) | 96.4 (94.8–97.7) | 93.5 | 95.5 | 96.4 | 94.0 | 0 / 0 | −37.4 (−41.7 to −33.2) | worse |
| Claude Sonnet 5 | 93.4 (91.4–95.2) | 98.0 (96.8–99.1) | 90.9 | 96.5 | 91.6 | 92.8 | 0 / 0 | −39.0 (−43.0 to −35.1) | worse |
| Gemini 3.5 Flash-Lite | 71.7 (68.1–75.2) | 77.7 (74.3–80.9) | 68.6 | 72.3 | 83.1 | 80.7 | 0 / 0 | −17.3 (−21.4 to −13.0) | worse |
| MedGemma 27B | 56.1 (52.4–59.9) | 66.0 (61.9–69.7) | 50.7 | 63.9 | 80.7 | 83.1 | 0 / 0 | −1.8 (−6.2 to +2.7) | inconclusive |
| Gemma 3 27B | 42.6 (39.0–46.0) | 48.1 (44.4–51.5) | 50.4 | 56.0 | 81.9 | 81.9 | 0 / 0 | +11.8 (+7.5 to +15.9) | non-inferior |

Highest KDIGO stage by the creatinine criteria in the first 72 hours, 561 eICU demo stays, one per patient: no kidney injury 250, stage 1 228, stage 2 33, stage 3 50. The sample is enriched for kidney injury, so overall accuracy does not reflect how common each stage is. Exact accuracy unless stated; balanced accuracy is the mean of the four stages' recall. Jev minus each system: paired bootstrap stratified by stage, Holm's adjustment across the ten LLMs, margin 5 points. Three or more of the five flagship LLMs agreed on a stage other than the key for 4 stays with the stage names only and for none with the KDIGO criteria.

### Supplementary Table 4. Cost and response time

List cost per 1,000 answers, US$:

| System | Risk of death | Patient questions | Board questions | Cancer staging, stage names | Cancer staging, medical context | Kidney-injury staging, stage names | Kidney-injury staging, KDIGO criteria |
|---|---|---|---|---|---|---|---|
| Jev 1.13 | 0.040 | 0.021 | 0.023 | 0.028 | 0.034 | 0.020 | 0.027 |
| GPT-5.6 Sol | 9.31 | 4.02 | 5.78 | 9.61 | 8.98 | 6.49 | 6.17 |
| Claude Opus 5.5 | 6.09 | 4.75 | 5.75 | 8.19 | 8.64 | 6.81 | 6.75 |
| Gemini 3.8 Flash | 2.69 | 1.56 | 1.96 | 2.62 | 2.47 | 3.15 | 2.55 |
| Muse Spark 1.1 | 4.88 | 2.86 | 3.56 | 5.69 | 5.86 | 5.91 | 6.10 |
| GLM-5.3 | 15.08 | 3.13 | 7.80 | 10.11 | 7.35 | 11.91 | 8.28 |
| GPT-5.6 Luna | 0.333 | 0.179 | 0.261 | 0.369 | 0.342 | 0.303 | 0.361 |
| Claude Sonnet 5 | 3.16 | 2.72 | 2.71 | 4.65 | 4.74 | 4.84 | 5.29 |
| Gemini 3.5 Flash-Lite | 0.307 | 0.310 | 0.307 | 0.600 | 0.625 | 0.340 | 0.393 |
| MedGemma 27B | 0.237 | 0.105 | 0.112 | 0.180 | 0.220 | 0.112 | 0.157 |
| Gemma 3 27B | 0.090 | 0.042 | 0.044 | 0.075 | 0.091 | 0.044 | 0.062 |
| Jev 1.13, recommended form | 0.035 | — | — | 0.117 | 0.133† | — | — |

Seconds from request to complete reply, median (90th percentile):

| System | Risk of death | Patient questions | Board questions | Cancer staging |
|---|---|---|---|---|
| Jev 1.13 | 0.96 (1.03) | 0.90 (1.09) | 0.94 (1.03) | 1.02 (1.13) |
| GPT-5.6 Sol | 4.66 (5.77) | 2.48 (3.77) | 3.66 (5.76) | 4.14 (5.52) |
| Claude Opus 5.5 | 2.52 (5.57) | 2.56 (4.86) | 2.69 (6.30) | 5.03 (7.17) |
| Gemini 3.8 Flash | 6.22 (7.56) | 4.68 (6.26) | 4.80 (7.64) | 5.20 (7.05) |
| Muse Spark 1.1 | 5.25 (6.72) | 3.07 (3.94) | 4.10 (7.08) | 5.48 (7.95) |
| GLM-5.3 | 36.81 (102.29) | 8.94 (20.19) | 11.18 (68.94) | 12.05 (30.25) |
| GPT-5.6 Luna | 2.94 (3.83) | 2.82 (3.44) | 3.12 (5.02) | 3.07 (3.85) |
| Claude Sonnet 5 | 2.41 (6.30) | 3.21 (5.34) | 2.66 (6.55) | 4.01 (6.67) |
| Gemini 3.5 Flash-Lite | 1.48 (1.79) | 1.54 (1.98) | 1.56 (1.77) | 1.80 (2.07) |
| MedGemma 27B | 3.76 (15.74) | 4.41 (16.60) | 4.68 (15.64) | 8.03 (21.91) |
| Gemma 3 27B | 7.50 (18.22) | 7.91 (19.93) | 9.06 (19.06) | 10.28 (28.72) |
| Jev 1.13, recommended form | 0.66 (0.82) | — | — | 0.79 (0.89) |

Cancer staging, recommended-form pairing on the 240 reports: estimated time as a share of the LLM alone:

| LLM | Pairing time, % of the LLM alone |
|---|---|
| GPT-5.6 Sol | 70 |
| Claude Opus 5.5 | 67 |
| Gemini 3.8 Flash | 66 |
| Muse Spark 1.1 | 65 |
| GLM-5.3 | 55 |

Cost: tokens at the standard list price in September 2026 (Supplementary Table 8); cancer staging on the 240 reports. †With the AJCC notes. Time: 100 items per system and task sent one at a time, all systems of a task timed in the same session; kidney-injury staging was not timed. Each model was timed alone; in the pairing, a report passed on waits for both answers, so its time is Jev's plus the LLM's, and a report Jev keeps takes Jev's alone; the pairing's share applies these times to the reports Jev kept and passed on among the 240. Tokens per answer and cost per correct answer are in the code repository.

### Supplementary Table 5. Cancer staging: every form of question, every system (240 reports)

#### a. Accuracy, % (95% CI)

| System | Stage names | Medical context | Task decomposition only | Both, options defined without examples | Both: recommended form | Recommended form with AJCC notes |
|---|---|---|---|---|---|---|
| Jev 1.13 | 45.4 (39.2–51.7) | 75.8 (70.0–80.8) | 82.1 (76.7–86.4) | 87.1 (82.2–90.7) | 85.8 (80.9–89.7) | 97.5 (94.7–98.8) |
| GPT-5.6 Sol | 99.2 (97.0–99.8) | 99.6 (97.7–99.9) | 99.6 (97.7–99.9) | 100.0 | 100.0 | 100.0 |
| Claude Opus 5.5 | 97.1 (94.1–98.6) | 100.0 | 99.6 (97.7–99.9) | 100.0 | 100.0 | 99.6 (97.7–99.9) |
| Gemini 3.8 Flash | 100.0 | 100.0 | 100.0 | 100.0 | 100.0 | 100.0 |
| Muse Spark 1.1 | 99.6 (97.7–99.9) | 100.0 | 99.6 (97.7–99.9) | 100.0 | 100.0 | 100.0 |
| GLM-5.3 | 94.2 (90.4–96.5) | 97.5 (94.7–98.8) | 97.9 (95.2–99.1) | 100.0 | 98.8 (96.4–99.6) | 100.0 |
| GPT-5.6 Luna | 97.1 (94.1–98.6) | 96.7 (93.6–98.3) | 94.6 (91.0–96.8) | 100.0 | 100.0 | 100.0 |
| Claude Sonnet 5 | 97.5 (94.7–98.8) | 95.8 (92.5–97.7) | 99.6 (97.7–99.9) | 100.0 | 100.0 | 100.0 |
| Gemini 3.5 Flash-Lite | 59.2 (52.9–65.2) | 73.8 (67.8–78.9) | 87.9 (83.2–91.5) | 90.8 (86.5–93.9) | 83.3 (78.1–87.5) | 98.8 (96.4–99.6) |
| MedGemma 27B | 30.0 (24.6–36.1) | 39.2 (33.2–45.5) | 49.6 (43.3–55.9) | 80.4 (74.9–84.9) | 74.6 (68.7–79.7) | 72.9 (67.0–78.1) |
| Gemma 3 27B | 26.7 (21.5–32.6) | 25.8 (20.7–31.7) | 41.7 (35.6–48.0) | 71.2 (65.2–76.6) | 65.4 (59.2–71.1) | 64.2 (57.9–70.0) |
| Jev + GPT-5.6 Sol | 99.2 (97.0–99.8) | 99.6 (97.7–99.9) | 99.6 (97.7–99.9) | 100.0 | 100.0 | 100.0 |
| Jev + Claude Opus 5.5 | 97.1 (94.1–98.6) | 100.0 | 99.6 (97.7–99.9) | 100.0 | 100.0 | 99.6 (97.7–99.9) |
| Jev + Gemini 3.8 Flash | 100.0 | 100.0 | 100.0 | 100.0 | 100.0 | 100.0 |
| Jev + Muse Spark 1.1 | 99.6 (97.7–99.9) | 100.0 | 99.6 (97.7–99.9) | 100.0 | 100.0 | 100.0 |
| Jev + GLM-5.3 | 94.2 (90.4–96.5) | 97.5 (94.7–98.8) | 97.9 (95.2–99.1) | 100.0 | 98.8 (96.4–99.6) | 100.0 |
| Jev + GPT-5.6 Luna | 97.1 (94.1–98.6) | 96.7 (93.6–98.3) | 94.6 (91.0–96.8) | 100.0 | 100.0 | 100.0 |
| Jev + Claude Sonnet 5 | 97.5 (94.7–98.8) | 96.2 (93.0–98.0) | 99.6 (97.7–99.9) | 100.0 | 100.0 | 100.0 |
| Jev + Gemini 3.5 Flash-Lite | 59.2 (52.9–65.2) | 73.8 (67.8–78.9) | 87.9 (83.2–91.5) | 90.8 (86.5–93.9) | 83.3 (78.1–87.5) | 98.8 (96.4–99.6) |
| Jev + MedGemma 27B | 30.0 (24.6–36.1) | 40.0 (34.0–46.3) | 50.4 (44.1–56.7) | 81.7 (76.3–86.1) | 75.8 (70.0–80.8) | 75.0 (69.2–80.1) |
| Jev + Gemma 3 27B | 26.7 (21.5–32.6) | 26.7 (21.5–32.6) | 42.5 (36.4–48.8) | 73.8 (67.8–78.9) | 70.0 (63.9–75.4) | 71.2 (65.2–76.6) |

Exact accuracy on the 240 reports; Wilson 95% intervals; unparsable answers count as wrong. In each form every system received character-identical reports, questions and options; the LLMs also received a system message giving the reply format (Supplementary Note 1). Stage names and medical context: one question with the ten stage names, or with each stage's T, N and M categories. Task decomposition only: four questions (T category, involved regional nodes, tumour deposits, distant metastasis) with bare options. Both: the four questions with each option defined, without or with examples; the recommended form adds examples and sends the report as a named field (Supplementary Note 2). In every four-question form the stage group was computed in code from the four answers with the table that built the key, for every system. Jev + LLM: Jev's answer where it was fully confident, and the LLM's answer in the same form otherwise (Jev kept 0, 2, 3, 86, 116, 126 of the 240 reports in the six forms, in the order of the columns); the pairing of Jev in the recommended form with the LLM given stage names is in Supplementary Table 7a. Jev with stage names and the report sent as a named field: 46.2 (40.1–52.6); for the LLMs this request was character-identical to the stage-names request.

**b. Jev minus each LLM in the same form, points (95% CI)**

| LLM | Stage names | Medical context | Task decomposition only | Both, options defined without examples | Both: recommended form | Recommended form with AJCC notes |
|---|---|---|---|---|---|---|
| GPT-5.6 Sol | −53.8 (−60.0 to −47.5) | −23.8 (−29.2 to −18.3) | −17.5 (−22.1 to −12.9) | −12.9 (−16.7 to −9.2) | −14.2 (−17.9 to −10.4) | −2.5 (−4.2 to −0.8) |
| Claude Opus 5.5 | −51.7 (−58.3 to −45.4) | −24.2 (−29.6 to −18.8) | −17.5 (−22.1 to −12.9) | −12.9 (−16.7 to −9.2) | −14.2 (−17.9 to −10.4) | −2.1 (−4.2 to 0.0) |
| Gemini 3.8 Flash | −54.6 (−60.8 to −48.3) | −24.2 (−29.6 to −18.8) | −17.9 (−22.5 to −13.3) | −12.9 (−16.7 to −9.2) | −14.2 (−17.9 to −10.4) | −2.5 (−4.2 to −0.8) |
| Muse Spark 1.1 | −54.2 (−60.4 to −47.9) | −24.2 (−29.6 to −18.8) | −17.5 (−22.1 to −12.9) | −12.9 (−16.7 to −9.2) | −14.2 (−17.9 to −10.4) | −2.5 (−4.2 to −0.8) |
| GLM-5.3 | −48.8 (−55.4 to −41.7) | −21.7 (−27.5 to −16.2) | −15.8 (−20.8 to −11.2) | −12.9 (−16.7 to −9.2) | −12.9 (−16.7 to −9.2) | −2.5 (−4.2 to −0.8) |
| GPT-5.6 Luna | −51.7 (−58.3 to −44.6) | −20.8 (−26.7 to −14.6) | −12.5 (−17.9 to −7.1) | −12.9 (−16.7 to −9.2) | −14.2 (−17.9 to −10.4) | −2.5 (−4.2 to −0.8) |
| Claude Sonnet 5 | −52.1 (−59.2 to −45.4) | −20.0 (−25.8 to −14.2) | −17.5 (−22.1 to −12.9) | −12.9 (−16.7 to −9.2) | −14.2 (−17.9 to −10.4) | −2.5 (−4.2 to −0.8) |
| Gemini 3.5 Flash-Lite | −13.8 (−21.7 to −5.8) | +2.1 (−4.6 to +8.3) | −5.8 (−10.0 to −2.1) | −3.8 (−6.2 to −1.2) | +2.5 (+0.8 to +4.6) | −1.2 (−2.9 to +0.8) |
| MedGemma 27B | +15.4 (+7.9 to +22.9) | +36.7 (+28.7 to +44.6) | +32.5 (+26.7 to +38.8) | +6.7 (+4.2 to +9.6) | +11.2 (+7.5 to +15.4) | +24.6 (+20.0 to +29.2) |
| Gemma 3 27B | +18.8 (+11.2 to +25.8) | +50.0 (+42.9 to +57.1) | +40.4 (+34.2 to +47.1) | +15.8 (+11.7 to +20.4) | +20.4 (+15.4 to +25.4) | +33.3 (+27.9 to +38.8) |

2,000 paired bootstrap resamples stratified by report type. Negative: Jev less accurate.

**c. Answers at full confidence, % (wrong answers among them)**

| System | Stage names | Medical context | Task decomposition only | Both, options defined without examples | Both: recommended form | Recommended form with AJCC notes |
|---|---|---|---|---|---|---|
| Jev 1.13 | 0 (0) | 1 (0) | 1 (0) | 36 (0) | 48 (0) | 52 (0) |
| GPT-5.6 Sol | 4 (0) | 7 (0) | 7 (0) | 9 (0) | 18 (0) | 20 (0) |
| Claude Opus 5.5 | 0 (0) | 0 (0) | 0 (0) | 0 (0) | 0 (0) | 0 (0) |
| Gemini 3.8 Flash | 3 (0) | 4 (0) | 2 (0) | 6 (0) | 9 (0) | 7 (0) |
| Muse Spark 1.1 | 21 (0) | 20 (0) | 3 (0) | 17 (0) | 25 (0) | 25 (0) |
| GLM-5.3 | 2 (0) | 5 (0) | 2 (0) | 6 (0) | 3 (0) | 3 (0) |
| GPT-5.6 Luna | 33 (0) | 35 (1) | 32 (0) | 51 (0) | 49 (0) | 52 (0) |
| Claude Sonnet 5 | 0 (0) | 0 (0) | 0 (0) | 6 (0) | 8 (0) | 10 (0) |
| Gemini 3.5 Flash-Lite | 63 (59) | 92 (57) | 27 (10) | 55 (16) | 65 (29) | 62 (2) |
| MedGemma 27B | 51 (77) | 68 (97) | 83 (105) | 71 (34) | 78 (46) | 80 (52) |
| Gemma 3 27B | 0 (1) | 0 (0) | 0 (0) | 1 (1) | 0 (1) | 0 (1) |

Full confidence: a top probability of 1.00 on the one question, or on each of the four questions (their product 1.00). The distribution of confidence in each form is in Supplementary Table 6.

**d. Jev in every form, by report type**

| Form of question | 240 reports | Ordinary (120) | Built to mislead (120) | Misleading feature (100) | Metastatic node at an unusual site (20) |
|---|---|---|---|---|---|
| Standard LLM-style inputs: one question, stage names | 45.4 (0) | 44.2 (0) | 46.7 (0) | 43.0 (0) | 65.0 (0) |
| The same, report sent as a named field | 46.2 (0) | 48.3 (0) | 44.2 (0) | 40.0 (0) | 65.0 (0) |
| Medical context: each stage’s T, N and M categories | 75.8 (1) | 80.8 (2) | 70.8 (0) | 75.0 (0) | 50.0 (0) |
| Task decomposition: four questions, bare options | 82.1 (1) | 92.5 (2) | 71.7 (0) | 78.0 (0) | 40.0 (0) |
| Both, options defined without examples | 87.1 (36) | 100.0 (47) | 74.2 (25) | 80.0 (30) | 45.0 (0) |
| Both: recommended form | 85.8 (48) | 100.0 (63) | 71.7 (33) | 80.0 (40) | 30.0 (0) |
| Recommended form with AJCC notes | 97.5 (52) | 100.0 (65) | 95.0 (40) | 100.0 (48) | 70.0 (0) |
| Flagship LLMs, stage names | 94.2–100.0 | 95.0–100.0 | 93.3–100.0 | 92.0–100.0 | 100.0 |
| Flagship LLMs, recommended form | 98.8–100.0 | 100.0 | 97.5–100.0 | 98.0–100.0 | 95.0–100.0 |

Jev’s exact accuracy, %; in brackets, % of its answers at full confidence. None of Jev’s fully confident answers was wrong, in any form or set. Report types: Supplementary Table 3a and Note 4. The AJCC notes’ examples of non-regional nodes named the two sites used in the misleading-feature reports and no site of the unusual-site reports (Methods).

### Supplementary Table 6. Confidence and correctness

| System | Answers | Correct, % | Below 0.75: % of answers | Below 0.75: Correct, % | 0.75–0.99: % of answers | 0.75–0.99: Correct, % | Fully confident (1.00): Answers (% of all) | Fully confident (1.00): Correct, % |
|---|---|---|---|---|---|---|---|---|
| **Cancer staging, 240 reports** | | | | | | | | |
| *Stage names only (standard LLM-style inputs)* | | | | | | | | |
| Jev 1.13 | 240 | 45.4 | 98 | 44.7 | 2 | 80.0 | 0 (0) | — |
| Flagship LLMs (5, pooled) | 1,200 | 98.0 | 1 | 92.9 | 92 | 98.9 | 73 (6) | 100.0 |
| Smaller LLMs (5, pooled) | 1,200 | 62.1 | 19 | 28.5 | 52 | 74.9 | 354 (30) | 61.3 |
| *Medical context: each stage's T, N and M categories* | | | | | | | | |
| Jev 1.13 | 240 | 75.8 | 54 | 56.6 | 45 | 98.2 | 2 (1) | 100.0 |
| Flagship LLMs (5, pooled) | 1,200 | 99.4 | 1 | 100.0 | 92 | 99.9 | 84 (7) | 100.0 |
| Smaller LLMs (5, pooled) | 1,200 | 66.2 | 10 | 25.8 | 51 | 74.1 | 467 (39) | 66.8 |
| *Task decomposition: four questions, options not defined* | | | | | | | | |
| Jev 1.13 | 240 | 82.1 | 21 | 49.0 | 78 | 90.9 | 3 (1) | 100.0 |
| Flagship LLMs (5, pooled) | 1,200 | 99.3 | 4 | 98.0 | 92 | 99.9 | 35 (3) | 100.0 |
| Smaller LLMs (5, pooled) | 1,200 | 74.7 | 13 | 55.8 | 58 | 82.8 | 344 (29) | 66.6 |
| *Both, options defined without examples* | | | | | | | | |
| Jev 1.13 | 240 | 87.1 | 7 | 50.0 | 58 | 83.3 | 86 (36) | 100.0 |
| Flagship LLMs (5, pooled) | 1,200 | 100.0 | 2 | 100.0 | 90 | 100.0 | 93 (8) | 100.0 |
| Smaller LLMs (5, pooled) | 1,200 | 88.5 | 12 | 72.2 | 51 | 92.4 | 441 (37) | 88.4 |
| *Recommended form: four questions, options defined with examples* | | | | | | | | |
| Jev 1.13 | 240 | 85.8 | 4 | 70.0 | 48 | 72.8 | 116 (48) | 100.0 |
| Flagship LLMs (5, pooled) | 1,200 | 99.8 | 3 | 100.0 | 86 | 100.0 | 132 (11) | 100.0 |
| Smaller LLMs (5, pooled) | 1,200 | 84.7 | 13 | 63.8 | 47 | 90.7 | 480 (40) | 84.2 |
| *Recommended form with AJCC notes* | | | | | | | | |
| Jev 1.13 | 240 | 97.5 | 5 | 50.0 | 42 | 100.0 | 126 (52) | 100.0 |
| Flagship LLMs (5, pooled) | 1,200 | 99.9 | 1 | 90.9 | 88 | 100.0 | 131 (11) | 100.0 |
| Smaller LLMs (5, pooled) | 1,200 | 87.2 | 11 | 62.8 | 48 | 91.6 | 491 (41) | 88.8 |
| **Kidney-injury staging, 561 stays** | | | | | | | | |
| *Stage names only* | | | | | | | | |
| Jev 1.13 | 561 | 54.4 | 65 | 43.8 | 35 | 74.0 | 0 (0) | — |
| Flagship LLMs (5, pooled) | 2,805 | 96.0 | 9 | 88.4 | 88 | 98.6 | 17 (1) | 100.0 |
| Smaller LLMs (5, pooled) | 2,805 | 71.1 | 29 | 42.9 | 64 | 83.0 | 194 (7) | 82.0 |
| *With each stage's KDIGO criteria* | | | | | | | | |
| Jev 1.13 | 561 | 91.8 | 35 | 79.5 | 59 | 98.2 | 37 (7) | 100.0 |
| Flagship LLMs (5, pooled) | 2,805 | 99.5 | 1 | 100.0 | 91 | 99.9 | 211 (8) | 100.0 |
| Smaller LLMs (5, pooled) | 2,805 | 77.3 | 21 | 43.6 | 69 | 85.7 | 292 (10) | 88.7 |
| **Choosing Wisely patient questions, 180** | | | | | | | | |
| *Question as written* | | | | | | | | |
| Jev 1.13 | 180 | 85.0 | 15 | 48.1 | 30 | 77.8 | 99 (55) | 99.0 |
| Flagship LLMs (5, pooled) | 900 | 99.3 | 2 | 95.0 | 98 | 99.4 | 0 (0) | — |
| Smaller LLMs (5, pooled) | 900 | 90.8 | 20 | 81.2 | 80 | 93.2 | 2 (<1) | 100.0 |
| **Board questions, 1,571** | | | | | | | | |
| *Published answer options* | | | | | | | | |
| Jev 1.13 | 1,571 | 85.6 | 23 | 56.4 | 47 | 91.0 | 465 (30) | 99.6 |
| Flagship LLMs (5, pooled) | 7,855 | 94.1 | 8 | 66.1 | 91 | 97.1 | 24 (<1) | 100.0 |
| Smaller LLMs (5, pooled) | 7,855 | 80.4 | 28 | 63.6 | 72 | 86.8 | 55 (1) | 94.5 |
| *A different number of options* | | | | | | | | |
| Jev 1.13 | 1,571 | 83.5 | 26 | 50.5 | 48 | 92.0 | 420 (27) | 100.0 |
| Flagship LLMs (5, pooled) | 7,855 | 93.5 | 8 | 63.0 | 91 | 97.1 | 28 (<1) | 100.0 |
| Smaller LLMs (5, pooled) | 7,855 | 79.1 | 32 | 63.5 | 68 | 86.3 | 46 (1) | 95.7 |

Confidence is the probability a system gave its chosen answer (for Jev's four-question forms, the product of the four). Unparsable answers have no confidence and count as wrong, so band shares can add up to less than 100%. Pooled rows sum the five systems' answers. Across all tasks and forms, 1,351 of Jev's 1,354 fully confident answers were correct: the three exceptions were one patient question and two Medbullets questions on which 9 and 10 of the 10 other systems gave Jev's answer. In the recommended form, all 34 of Jev's wrong answers were short of full confidence, and 116 of its 206 correct answers reached it. Risk of death is not included, because its answer is a probability with no separate confidence. The five flagship LLMs one by one, stage names: 78–97% of answers at 0.75–0.99; each system's share at full confidence in every form is in Supplementary Table 5c.

### Supplementary Table 7. Jev with an LLM

#### a. Jev in the recommended form kept at full confidence, an LLM otherwise (cancer staging)

| LLM | Correct, pairing / LLM alone | Difference from the LLM alone, points (95% CI) | | | | Errors across stage 0–II, III or IV, pairing / LLM alone | Cost, % of the LLM-only cost |
|---|---|---|---|---|---|---|---|
| | | 240 reports | Ordinary (120) | Misleading feature (100) | Unusual node site (20) | | |
| **Recommended form. Kept by Jev: 240 reports: 116 of 240 (48%); ordinary: 76 of 120 (63%); misleading feature: 40 of 100 (40%); unusual node site: 0 of 20 (0%). None of the kept answers was wrong.** | | | | | | | |
| GPT-5.6 Sol | 238 / 238 | 0.0 | 0.0 | 0.0 | 0.0 | 0 / 0 | 56 |
| Claude Opus 5.5 | 236 / 233 | +1.2 (0.0 to +2.9) | +2.5 (0.0 to +5.8) | 0.0 | 0.0 | 0 / 0 | 56 |
| Gemini 3.8 Flash | 240 / 240 | 0.0 | 0.0 | 0.0 | 0.0 | 0 / 0 | 60 |
| Muse Spark 1.1 | 239 / 239 | 0.0 | 0.0 | 0.0 | 0.0 | 0 / 0 | 58 |
| GLM-5.3 | 230 / 226 | +1.7 (+0.4 to +3.3) | +2.5 (0.0 to +5.8) | +1.0 (0.0 to +3.0) | 0.0 | 8 / 11 | 67 |
| GPT-5.6 Luna | 233 / 233 | 0.0 | 0.0 | 0.0 | 0.0 | 0 / 0 | 89 |
| Claude Sonnet 5 | 236 / 234 | +0.8 (0.0 to +2.1) | +1.7 (0.0 to +4.2) | 0.0 | 0.0 | 0 / 0 | 58 |
| Gemini 3.5 Flash-Lite | 193 / 142 | +21.2 (+16.7 to +26.7) | +30.8 (+23.3 to +39.2) | +14.0 (+8.0 to +21.0) | 0.0 | 3 / 7 | 71 |
| MedGemma 27B | 171 / 72 | +41.2 (+35.4 to +47.5) | +53.3 (+45.0 to +62.5) | +35.0 (+25.0 to +44.0) | 0.0 | 41 / 96 | 116 |
| Gemma 3 27B | 164 / 64 | +41.7 (+35.8 to +47.5) | +53.3 (+45.0 to +62.5) | +36.0 (+27.0 to +45.0) | 0.0 | 49 / 97 | 207 |
| **Recommended form with AJCC notes. Kept by Jev: 240 reports: 126 of 240 (52%); ordinary: 78 of 120 (65%); misleading feature: 48 of 100 (48%); unusual node site: 0 of 20 (0%). None of the kept answers was wrong.** | | | | | | | |
| GPT-5.6 Sol | 238 / 238 | 0.0 | 0.0 | 0.0 | 0.0 | 0 / 0 | 53 |
| Claude Opus 5.5 | 236 / 233 | +1.2 (0.0 to +2.9) | +2.5 (0.0 to +5.8) | 0.0 | 0.0 | 0 / 0 | 52 |
| Gemini 3.8 Flash | 240 / 240 | 0.0 | 0.0 | 0.0 | 0.0 | 0 / 0 | 56 |
| Muse Spark 1.1 | 239 / 239 | 0.0 | 0.0 | 0.0 | 0.0 | 0 / 0 | 54 |
| GLM-5.3 | 230 / 226 | +1.7 (+0.4 to +3.3) | +2.5 (0.0 to +5.8) | +1.0 (0.0 to +3.0) | 0.0 | 8 / 11 | 64 |
| GPT-5.6 Luna | 233 / 233 | 0.0 | 0.0 | 0.0 | 0.0 | 0 / 0 | 89 |
| Claude Sonnet 5 | 236 / 234 | +0.8 (0.0 to +2.1) | +1.7 (0.0 to +4.2) | 0.0 | 0.0 | 0 / 0 | 54 |
| Gemini 3.5 Flash-Lite | 197 / 142 | +22.9 (+17.9 to +28.3) | +33.3 (+25.8 to +42.5) | +15.0 (+8.0 to +22.0) | 0.0 | 3 / 7 | 69 |
| MedGemma 27B | 182 / 72 | +45.8 (+39.6 to +52.1) | +56.7 (+48.3 to +65.0) | +42.0 (+32.0 to +52.0) | 0.0 | 36 / 96 | 121 |
| Gemma 3 27B | 172 / 64 | +45.0 (+39.2 to +51.2) | +54.2 (+45.0 to +62.5) | +43.0 (+33.0 to +52.0) | 0.0 | 46 / 97 | 224 |

Jev answered every report in the recommended form; its answer stood at full confidence (1.00 on all four questions) and the LLM's answer with stage names only was used otherwise. Correct answers, errors across groups and cost refer to the 240 reports. Difference: pairing minus the LLM alone, 2,000 paired bootstrap resamples stratified by report type; the lowest one-sided bound, Bonferroni-adjusted over the ten LLMs, across every set and LLM was 0.0 points (margin −5). An unparsable answer counts as wrong and as an error across groups. Cost at list price: Jev on every report and the LLM on the reports passed on. At a threshold of 0.99, 2 of Jev's wrong answers without the notes, and 0 with them, would have been kept. Report types: Supplementary Table 3a and Note 4.

With the LLM also in the recommended form (the reports Jev passed on answered by the LLM in the same four-question form; 240 reports):

| LLM | Correct, pairing / LLM alone | Difference, points (95% CI) | Errors across stage 0–II, III or IV, pairing / LLM alone | US$ per 1,000 reports, pairing / LLM alone | Cost, % of the LLM-only cost |
|---|---|---|---|---|---|
| GPT-5.6 Sol | 240 / 240 | 0.0 | 0 / 0 | 10.78 / 20.33 | 53 |
| Claude Opus 5.5 | 240 / 240 | 0.0 | 0 / 0 | 14.30 / 27.24 | 52 |
| Gemini 3.8 Flash | 240 / 240 | 0.0 | 0 / 0 | 2.97 / 5.32 | 56 |
| Muse Spark 1.1 | 240 / 240 | 0.0 | 0 / 0 | 3.95 / 7.18 | 55 |
| GLM-5.3 | 237 / 237 | 0.0 | 3 / 3 | 7.69 / 11.10 | 69 |
| GPT-5.6 Luna | 240 / 240 | 0.0 | 0 / 0 | 0.58 / 0.87 | 66 |
| Claude Sonnet 5 | 240 / 240 | 0.0 | 0 / 0 | 7.81 / 14.38 | 54 |
| Gemini 3.5 Flash-Lite | 200 / 200 | 0.0 | 39 / 39 | 1.17 / 2.05 | 57 |
| MedGemma 27B | 182 / 179 | +1.2 (0.0 to +2.9) | 40 / 40 | 0.58 / 0.91 | 64 |
| Gemma 3 27B | 168 / 157 | +4.6 (+2.1 to +7.5) | 40 / 40 | 0.31 / 0.37 | 83 |

Jev kept the same 116 reports; the LLM alone is the same LLM in the recommended form. Paired bootstrap stratified by report type (2,000 resamples).

#### b. Jev kept at full confidence, an LLM otherwise: the other tasks (every system received the same text)

| LLM | Correct, %, pairing / LLM alone | Difference, points (95% CI) | Adjusted lower bound, points | Cost, % of the LLM-only cost | US$ per 1,000 answers, pairing / LLM alone |
|---|---|---|---|---|---|
| **Patient questions (balanced accuracy). Kept by Jev: 99 of 180 (55%), 1 of them wrong.** | | | | | |
| GPT-5.6 Sol | 99.6 / 100.0 | −0.4 (−1.2 to 0.0) | −2.0 | 53 | 2.12 / 4.02 |
| Claude Opus 5.5 | 99.6 / 100.0 | −0.4 (−1.2 to 0.0) | −2.0 | 52 | 2.46 / 4.75 |
| Gemini 3.8 Flash | 99.6 / 99.6 | 0.0 | 0.0 | 53 | 0.82 / 1.56 |
| Muse Spark 1.1 | 98.7 / 99.1 | −0.4 (−1.2 to 0.0) | −2.0 | 49 | 1.39 / 2.86 |
| GLM-5.3 | 98.4 / 98.4 | 0.0 | 0.0 | 57 | 1.80 / 3.13 |
| GPT-5.6 Luna | 99.2 / 99.6 | −0.4 (−1.2 to 0.0) | −2.0 | 62 | 0.11 / 0.18 |
| Claude Sonnet 5 | 97.5 / 97.5 | 0.0 | 0.0 | 54 | 1.47 / 2.72 |
| Gemini 3.5 Flash-Lite | 95.5 / 95.5 | 0.0 | 0.0 | 52 | 0.16 / 0.31 |
| MedGemma 27B | 86.1 / 86.1 | 0.0 | 0.0 | 65 | 0.07 / 0.11 |
| Gemma 3 27B | 85.0 / 85.0 | 0.0 | 0.0 | 95 | 0.040 / 0.042 |
| **Board questions. Kept by Jev: 465 of 1,571 (30%), 2 of them wrong.** | | | | | |
| GPT-5.6 Sol | 94.7 / 94.7 | 0.0 | 0.0 | 80 | 4.62 / 5.78 |
| Claude Opus 5.5 | 95.9 / 95.9 | +0.1 (0.0 to +0.2) | 0.0 | 79 | 4.53 / 5.75 |
| Gemini 3.8 Flash | 95.7 / 95.7 | 0.0 | 0.0 | 81 | 1.58 / 1.96 |
| Muse Spark 1.1 | 92.8 / 92.8 | 0.0 | 0.0 | 78 | 2.77 / 3.56 |
| GLM-5.3 | 91.3 / 91.2 | +0.1 (−0.1 to +0.3) | −0.3 | 90 | 7.03 / 7.80 |
| GPT-5.6 Luna | 91.9 / 91.9 | 0.0 | 0.0 | 88 | 0.23 / 0.26 |
| Claude Sonnet 5 | 92.2 / 92.2 | 0.0 | 0.0 | 78 | 2.12 / 2.71 |
| Gemini 3.5 Flash-Lite | 86.2 / 86.1 | +0.1 (0.0 to +0.3) | 0.0 | 79 | 0.24 / 0.31 |
| MedGemma 27B | 69.9 / 68.2 | +1.7 (+1.1 to +2.4) | +0.9 | 92 | 0.10 / 0.11 |
| Gemma 3 27B | 66.1 / 63.4 | +2.7 (+2.0 to +3.6) | +1.7 | 123 | 0.055 / 0.044 |

| LLM | Correct, %, pairing / LLM alone | Difference, points (95% CI) | Adjusted lower bound, points | Cost, % of the LLM-only cost | US$ per 1,000 answers, pairing / LLM alone |
|---|---|---|---|---|---|
| **Kidney-injury staging, with KDIGO criteria. Kept by Jev: 37 of 561 (7%), 0 of them wrong.** | | | | | |
| GPT-5.6 Sol | 100.0 / 100.0 | 0.0 | 0.0 | 96 | 5.92 / 6.17 |
| Claude Opus 5.5 | 99.8 / 99.8 | 0.0 | 0.0 | 95 | 6.44 / 6.75 |
| Gemini 3.8 Flash | 100.0 / 100.0 | 0.0 | 0.0 | 97 | 2.48 / 2.55 |
| Muse Spark 1.1 | 100.0 / 100.0 | 0.0 | 0.0 | 96 | 5.84 / 6.10 |
| GLM-5.3 | 97.9 / 97.9 | 0.0 | 0.0 | 97 | 8.04 / 8.28 |
| GPT-5.6 Luna | 96.4 / 96.4 | 0.0 | 0.0 | 103 | 0.37 / 0.36 |
| Claude Sonnet 5 | 98.0 / 98.0 | 0.0 | 0.0 | 97 | 5.14 / 5.29 |
| Gemini 3.5 Flash-Lite | 77.7 / 77.7 | 0.0 | 0.0 | 100 | 0.39 / 0.39 |
| MedGemma 27B | 66.1 / 66.0 | +0.2 (0.0 to +0.5) | 0.0 | 111 | 0.17 / 0.16 |
| Gemma 3 27B | 50.3 / 48.1 | +2.1 (+1.1 to +3.4) | +0.7 | 137 | 0.085 / 0.062 |

Jev answered every item and its answer stood at full confidence (1.00); the LLM answered the rest. Difference: pairing minus the LLM alone, 2,000 paired bootstrap resamples stratified by question type (patient questions) or stage level (kidney-injury staging). Adjusted lower bound: one-sided, Bonferroni over the ten LLMs; non-inferior when above −5 points (every row). Balanced accuracy for patient questions, exact accuracy otherwise. Cost at the list prices of Supplementary Table 8: Jev on every item and the LLM on the items passed on; with the cheapest models, Jev's own cost can exceed what it saves (above 100%). Cancer staging: panel a.

#### c. Every confidence threshold: accuracy, cost and money

| | | Difference from the LLM alone, points | | | | Cost, % of the LLM-only cost | | | |
|---|---|---|---|---|---|---|---|---|---|
| Confidence threshold | Jev answers, % | Flagship LLMs | Smaller LLMs | Adjusted lower bound, points (ten LLMs) | Non-inferior to all ten LLMs | Flagship LLMs | Smaller LLMs | Saved, US$ per 1,000 answers (flagship LLMs) | Added errors per 1,000 answers (flagship LLMs) |
| **Patient questions (balanced accuracy)** | | | | | | | | | |
| 1.00 (full confidence) | 55 | −0.4 to 0.0 | −0.4 to 0.0 | −2.0 | Yes | 49–57 | 52–95 | 0.74–2.29 | 0.0 to +5.6 |
| 0.99 (lowest non-inferior) | 63 | −2.0 to −0.8 | −2.0 to 0.0 | −4.8 | Yes | 40–46 | 44–87 | 0.89–2.75 | +11.1 to +27.8 |
| 0.95 | 71 | −2.8 to −1.6 | −2.8 to 0.0 | −6.0 | No | 32–38 | 35–79 | 1.02–3.14 | +22.2 to +38.9 |
| 0.90 | 76 | −4.0 to −2.8 | −4.0 to 0.0 | −7.6 | No | 27–33 | 30–74 | 1.11–3.42 | +38.9 to +55.6 |
| 0.80 | 83 | −5.2 to −3.6 | −4.8 to +1.2 | −9.2 | No | 19–23 | 22–67 | 1.26–3.73 | +50.0 to +72.2 |
| 0.70 | 88 | −6.9 to −5.3 | −6.5 to +1.6 | −12.2 | No | 13–17 | 15–62 | 1.36–4.00 | +66.7 to +88.9 |
| **Board questions** | | | | | | | | | |
| 1.00 (full confidence) | 30 | 0.0 to +0.1 | 0.0 to +2.7 | −0.3 | Yes | 78–90 | 78–123 | 0.38–1.22 | −0.6 to 0.0 |
| 0.99 | 38 | 0.0 to +0.2 | 0.0 to +4.6 | −0.2 | Yes | 69–86 | 71–115 | 0.50–1.78 | −1.9 to 0.0 |
| 0.95 | 54 | −0.3 to +0.1 | −0.3 to +9.1 | −0.9 | Yes | 53–75 | 55–99 | 0.78–2.73 | −1.3 to +3.2 |
| 0.90 | 62 | −0.7 to +0.1 | −0.3 to +11.2 | −1.5 | Yes | 45–67 | 46–91 | 0.94–3.15 | −0.6 to +7.0 |
| 0.80 | 72 | −1.8 to −0.4 | −1.3 to +14.4 | −3.3 | Yes | 32–54 | 36–80 | 1.16–3.92 | +4.5 to +18.5 |
| 0.72 (lowest non-inferior) | 80 | −3.1 to −1.1 | −1.9 to +16.9 | −4.8 | Yes | 24–42 | 28–73 | 1.34–4.50 | +10.8 to +31.2 |
| 0.70 | 81 | −3.4 to −1.3 | −2.1 to +17.4 | −5.1 | No | 23–39 | 27–71 | 1.37–4.75 | +13.4 to +33.7 |
| **Cancer staging, recommended form (240 reports)** | | | | | | | | | |
| 1.00 (full confidence) | 48 | 0.0 to +1.7 | 0.0 to +41.7 | 0.0 | Yes | 56–67 | 58–207 | 1.06–4.19 | −16.7 to 0.0 |
| 0.99 | 68 | −0.8 to +1.7 | −0.8 to +56.7 | −2.5 | Yes | 36–50 | 39–187 | 1.56–6.11 | −16.7 to +8.3 |

| Confidence threshold | Jev answers, % | Difference from the LLM alone, points: Flagship LLMs | Difference from the LLM alone, points: Smaller LLMs | Adjusted lower bound, points (ten LLMs) | Non-inferior to all ten LLMs | Cost, % of the LLM-only cost: Flagship LLMs | Cost, % of the LLM-only cost: Smaller LLMs | Saved, US$ per 1,000 answers (flagship LLMs) | Added errors per 1,000 answers (flagship LLMs) |
|---|---|---|---|---|---|---|---|---|---|
| 0.97 (lowest non-inferior) | 78 | −1.3 to +2.9 | −0.4 to +63.3 | −3.3 | Yes | 26–31 | 28–178 | 1.87–7.07 | −29.2 to +12.5 |
| 0.95 | 87 | −4.2 to +1.3 | −1.7 to +65.0 | −7.9 | No | 16–18 | 19–169 | 2.16–8.49 | −12.5 to +41.7 |
| 0.90 | 91 | −7.9 to −2.5 | −5.4 to +62.9 | −12.9 | No | 11–13 | 14–165 | 2.28–8.85 | +25.0 to +79.2 |
| 0.80 | 94 | −10.8 to −5.4 | −8.3 to +60.8 | −15.8 | No | 8–10 | 10–162 | 2.35–9.06 | +54.2 to +108.3 |
| 0.70 | 96 | −12.9 to −7.5 | −10.4 to +59.2 | −18.3 | No | 5–8 | 7–159 | 2.42–9.36 | +75.0 to +129.2 |
| **Kidney-injury staging, with KDIGO criteria** | | | | | | | | | |
| 1.00 (full confidence) | 7 | 0.0 | 0.0 to +2.1 | 0.0 | Yes | 95–97 | 97–137 | 0.07–0.31 | 0.0 |
| 0.99 | 14 | 0.0 | 0.0 to +6.8 | 0.0 | Yes | 90–93 | 93–130 | 0.19–0.69 | 0.0 |
| 0.95 | 24 | −0.2 to 0.0 | −0.2 to +11.9 | −0.9 | Yes | 81–86 | 83–120 | 0.38–1.24 | 0.0 to +1.8 |
| 0.90 | 36 | −0.2 to 0.0 | −0.2 to +18.0 | −0.9 | Yes | 70–77 | 71–108 | 0.66–2.02 | 0.0 to +1.8 |
| 0.80 | 58 | −0.4 to +0.2 | −0.2 to +27.5 | −1.2 | Yes | 47–55 | 49–86 | 1.26–3.71 | −1.8 to +3.6 |
| 0.70 | 73 | −1.8 to −0.9 | −1.2 to +34.0 | −3.4 | Yes | 31–39 | 34–71 | 1.67–5.04 | +8.9 to +17.8 |
| 0.63 (lowest non-inferior) | 80 | −2.7 to −1.4 | −2.0 to +37.1 | −4.6 | Yes | 23–29 | 25–64 | 1.89–5.90 | +14.3 to +26.7 |

Jev answers the items at or above the threshold and the LLM the rest. Thresholds were examined descriptively with the non-inferiority test of the main analysis, whose threshold was full confidence (margin 5 points; 2,000 paired bootstrap resamples; one-sided lower bound over all ten LLMs, Bonferroni over the ten, not over thresholds). Saved: the LLM-only cost minus the pairing's cost, at the list prices of Supplementary Table 8. Added errors: wrong answers of the pairing minus those of the LLM alone, per 1,000 items (negative: fewer errors than the LLM alone); raw counts for patient questions, whose difference column is in balanced accuracy. Cancer staging: the 240 reports, Jev in the recommended form, the LLMs with stage names. Risk of death is not included, because its answer is a probability of death, with no separate confidence.

### Supplementary Table 8. Systems tested

| System | Developer | Consumer plan, 25 September 2026 | Model identifier | Provider | List price, US$ per million tokens (input / output) | Training cutoff stated by the developer |
|---|---|---|---|---|---|---|
| Jev 1.13 | TypeSafe AI | — | typesafe/jev-1.13-20260917 | TypeSafe | 0.042 / 0 | Not published |
| GPT-5.6 Sol | OpenAI | ChatGPT Plus | openai/gpt-5.6-sol-20260709 | OpenAI | 5.00 / 30.00 | February 2026 |
| Claude Opus 5.5 | Anthropic | Claude Pro | anthropic/claude-opus-5.5-20260921 | Anthropic | 4.00 / 20.00 | June 2026 |
| Gemini 3.8 Flash | Google | Google AI Pro | google/gemini-3.8-flash-20260902 | Google Vertex AI | 0.75 / 3.75 | March 2026* |
| Muse Spark 1.1 | Meta | Meta AI app | meta/muse-spark-1.1-20260709 | Meta | 1.25 / 4.25 | Not published |
| GLM-5.3 | Z.ai | GLM Coding Plan | z-ai/glm-5.3-20260816 | Z.ai | 1.40 / 4.40 | Not published |
| GPT-5.6 Luna | OpenAI | ChatGPT Free | openai/gpt-5.6-luna-20260709 | OpenAI | 0.20 / 1.20 | February 2026 |
| Claude Sonnet 5 | Anthropic | Claude Free | anthropic/claude-sonnet-5-20260630 | Anthropic | 2.00 / 10.00 | January 2026 |
| Gemini 3.5 Flash-Lite | Google | Gemini app (free) | google/gemini-3.5-flash-lite-20260721 | Google Vertex AI | 0.30 / 2.50 | March 2026* |
| MedGemma 27B (text) | Google | — (open weights) | google/medgemma-27b-text-it | Featherless (8-bit weights), via the Hugging Face router | 0.265 / 0.65 | Not published |
| Gemma 3 27B | Google | — (open weights) | google/gemma-3-27b-it | Featherless (8-bit weights), via the Hugging Face router | 0.10 / 0.30 | August 2024 |

The five flagship LLMs listed after Jev form the main comparison; every non-inferiority test covers all ten LLMs. All calls went through OpenRouter to the pinned provider with fallbacks disabled, except MedGemma and Gemma. Every cost in this paper is at the standard list price; for GPT-5.6 Luna, the price on OpenAI's API price list[2]. Default settings throughout (no temperature or reasoning parameter; output capped at 16,000 tokens; seed 20260922 where accepted). Calls were made between 26 and 30 September 2026. Every stored answer reported the pinned provider and model, except that the Hugging Face router reports no provider and a few GLM-5.3 replies carried none.

*The developer states March 2026, with knowledge in some domains limited to January 2025.

### Supplementary Table 9. Ten-year risk: cohort and participants

| Step | n |
|---|---|
| NHANES respondents, 1999–2008 | 51,623 |
| Aged 40–79 at examination | 14,737 |
| Eligible for mortality linkage | 14,721 |
| Follow-up time from examination available | 14,008 |
| At least seven of nine routine blood biomarkers* | 13,171 |
| Vital status undetermined at 120 months | 0 |
| Analysis cohort (deaths within 120 months, 2,093) | 13,171 |
| Fitting set (reference model) | 11,971 |
| Software-test set | 200 |
| Evaluation set (all nine biomarkers) | 1,000 |

*Albumin, creatinine, glucose, C-reactive protein, lymphocyte percentage, mean cell volume, red cell distribution width, alkaline phosphatase and white cell count. Software-test and evaluation participants had all nine.

Baseline prediction used the 12,910 records with all nine biomarkers outside the software-test set.

Characteristics of the fitting and evaluation sets:

| Characteristic | Fitting set (n = 11,971) | Evaluation set (n = 1,000) |
|---|---|---|
| Survey cycle, n (%) | | |
| 1999–2000 | 2,137 (17.9%) | 187 (18.7%) |
| 2001–2002 | 2,397 (20.0%) | 192 (19.2%) |
| 2003–2004 | 2,239 (18.7%) | 215 (21.5%) |
| 2005–2006 | 2,223 (18.6%) | 180 (18.0%) |
| 2007–2008 | 2,975 (24.9%) | 226 (22.6%) |
| Age, years, median (IQR) | 58 (48–67) | 59 (48–67) |
| Sex, n (%) | | |
| Male | 6,000 (50.1%) | 519 (51.9%) |
| Female | 5,971 (49.9%) | 481 (48.1%) |
| Race and ethnicity, n (%) | | |
| Non-Hispanic White | 6,009 (50.2%) | 516 (51.6%) |
| Non-Hispanic Black | 2,462 (20.6%) | 181 (18.1%) |
| Mexican American | 2,432 (20.3%) | 212 (21.2%) |
| Other Hispanic | 660 (5.5%) | 59 (5.9%) |
| Other race including multiracial | 408 (3.4%) | 32 (3.2%) |
| Education, n (%) | | |
| Less than 9th grade | 2,042 (17.1%) | 172 (17.2%) |
| 9th to 11th grade | 1,898 (15.9%) | 153 (15.3%) |
| High school graduate or GED | 2,799 (23.4%) | 219 (21.9%) |
| Some college | 2,919 (24.4%) | 250 (25.0%) |
| College graduate or above | 2,299 (19.2%) | 204 (20.4%) |
| Missing | 14 (0.1%) | 2 (0.2%) |
| Family income to poverty ratio, median (IQR) | 2.48 (1.24–4.54) | 2.49 (1.24–4.55) |
| Missing, n (%) | 962 (8.0%) | 80 (8.0%) |
| Health insurance, n (%) | | |
| Yes | 9,997 (83.5%) | 826 (82.6%) |
| No | 1,893 (15.8%) | 169 (16.9%) |
| Missing | 81 (0.7%) | 5 (0.5%) |
| Smoking, n (%) | | |
| Never | 5,609 (46.9%) | 452 (45.2%) |
| Former | 3,796 (31.7%) | 340 (34.0%) |
| Current | 2,550 (21.3%) | 208 (20.8%) |
| Missing | 16 (0.1%) | 0 (0.0%) |
| Self-rated health, n (%) | | |
| Excellent | 1,667 (13.9%) | 142 (14.2%) |
| Very good | 2,968 (24.8%) | 229 (22.9%) |
| Good | 3,988 (33.3%) | 343 (34.3%) |
| Fair | 2,570 (21.5%) | 221 (22.1%) |
| Poor | 771 (6.4%) | 63 (6.3%) |
| Missing | 7 (0.1%) | 2 (0.2%) |
| Body mass index, kg/m², median (IQR) | 28.3 (24.9–32.3) | 28.4 (25.4–32.4) |

| Characteristic | Fitting set (n = 11,971) | Evaluation set (n = 1,000) |
|---|---|---|
| Missing, n (%) | 260 (2.2%) | 24 (2.4%) |
| Waist circumference, cm, median (IQR) | 99.4 (90.2–109.3) | 100.2 (91.5–109.5) |
| Missing, n (%) | 448 (3.7%) | 32 (3.2%) |
| Systolic blood pressure, mmHg, median (IQR) | 126 (115–140) | 127 (116–140) |
| Missing, n (%) | 462 (3.9%) | 46 (4.6%) |
| Diastolic blood pressure, mmHg, median (IQR) | 73 (65–80) | 73 (66–80) |
| Missing, n (%) | 462 (3.9%) | 46 (4.6%) |
| Prescription medicines in the past 30 days, median (IQR) | 2 (0–4) | 2 (0–4) |
| Diabetes, n (%) | | |
| Yes | 1,810 (15.1%) | 155 (15.5%) |
| Borderline | 238 (2.0%) | 17 (1.7%) |
| Missing | 7 (0.1%) | 1 (0.1%) |
| Congestive heart failure, n (%) | 496 (4.1%) | 35 (3.5%) |
| Missing | 55 (0.5%) | 4 (0.4%) |
| Coronary heart disease, n (%) | 665 (5.6%) | 64 (6.4%) |
| Missing | 71 (0.6%) | 5 (0.5%) |
| Myocardial infarction, n (%) | 713 (6.0%) | 51 (5.1%) |
| Missing | 26 (0.2%) | 0 (0.0%) |
| Stroke, n (%) | 537 (4.5%) | 48 (4.8%) |
| Missing | 24 (0.2%) | 2 (0.2%) |
| Emphysema, n (%) | 347 (2.9%) | 19 (1.9%) |
| Missing | 24 (0.2%) | 2 (0.2%) |
| Chronic bronchitis, n (%) | 875 (7.3%) | 71 (7.1%) |
| Missing | 29 (0.2%) | 1 (0.1%) |
| Cancer, n (%) | 1,296 (10.8%) | 93 (9.3%) |
| Missing | 16 (0.1%) | 3 (0.3%) |
| Serum albumin, g/dL, median (IQR) | 4.2 (4.0–4.4) | 4.2 (4.0–4.4) |
| Serum creatinine, mg/dL, median (IQR) | 0.86 (0.75–1.02) | 0.90 (0.75–1.06) |
| Serum glucose (random), mg/dL, median (IQR) | 94 (87–105) | 94 (87–104) |
| Missing, n (%) | 1 (0.0%) | 0 (0.0%) |
| C-reactive protein, mg/dL, median (IQR) | 0.24 (0.10–0.55) | 0.24 (0.10–0.52) |
| Missing, n (%) | 8 (0.1%) | 0 (0.0%) |
| Lymphocytes, %, median (IQR) | 29.8 (24.5–35.3) | 29.6 (24.4–36.0) |
| Missing, n (%) | 51 (0.4%) | 0 (0.0%) |
| Mean corpuscular volume, fL, median (IQR) | 90.5 (87.3–93.5) | 90.6 (87.3–93.3) |
| Red cell distribution width, %, median (IQR) | 12.6 (12.2–13.3) | 12.6 (12.2–13.3) |
| Alkaline phosphatase, U/L, median (IQR) | 71 (58–87) | 71 (58–86) |
| Missing, n (%) | 1 (0.0%) | 0 (0.0%) |
| White blood cell count, $\times 10^3/\mu L$, median (IQR) | 6.9 (5.6–8.2) | 6.9 (5.8–8.2) |
| HbA1c, %, median (IQR) | 5.5 (5.3–5.9) | 5.5 (5.3–5.9) |
| Missing, n (%) | 16 (0.1%) | 1 (0.1%) |
| Total cholesterol, mg/dL, median (IQR) | 203 (178–231) | 206 (179–234) |
| Missing, n (%) | 12 (0.1%) | 0 (0.0%) |
| HDL cholesterol, mg/dL, median (IQR) | 50 (41–61) | 49 (40–61) |
| Missing, n (%) | 11 (0.1%) | 2 (0.2%) |
| Urine albumin to creatinine ratio, mg/g, median (IQR) | 7.3 (4.6–15.5) | 7.3 (4.6–16.6) |
| Missing, n (%) | 192 (1.6%) | 12 (1.2%) |
| Died within ten years, n (%) | 1,899 (15.9%) | 163 (16.3%) |

Unweighted counts of the NHANES 1999–2008 participants aged 40–79 with ten-year mortality follow-up; the references were fitted on the fitting set, and the systems answered the 12,910 records with all nine biomarkers. IQR, interquartile range; HDL, high-density lipoprotein; HbA1c, glycated haemoglobin.

### Supplementary Table 10. Ten-year risk: response to one-field edits

| System | Slope, spline reference (simultaneous 95% CI) | Slope, LightGBM reference (simultaneous 95% CI) | Without glycated haemoglobin +1.0 point | Without creatinine ×1.5 | Without albumin −0.5 g/dL | Without systolic pressure +20 mmHg | Without never to current smoker | Without age +5 years |
|---|---|---|---|---|---|---|---|---|
| Jev 1.13 | 0.37 (0.20–0.53) | 0.39 (0.22–0.56) | 0.37 | 0.35 | 0.38 | 0.35 | 0.81 | 0.23 |
| GPT-5.6 Sol | 0.99 (0.67–1.32) | 1.03 (0.72–1.35) | 0.99 | 0.98 | 1.02 | 0.99 | 1.17 | 0.92 |
| Claude Opus 5.5 | 0.96 (0.65–1.27) | 1.00 (0.71–1.28) | 0.96 | 0.95 | 0.99 | 0.96 | 1.02 | 0.92 |
| Gemini 3.8 Flash | 0.98 (0.66–1.29) | 1.02 (0.72–1.32) | 0.98 | 0.96 | 1.00 | 0.98 | 1.09 | 0.94 |
| Muse Spark 1.1 | 0.82 (0.53–1.11) | 0.86 (0.57–1.14) | 0.82 | 0.79 | 0.83 | 0.81 | 1.13 | 0.73 |
| GLM-5.3 | 1.03 (0.71–1.35) | 1.07 (0.76–1.37) | 1.03 | 1.04 | 1.06 | 1.03 | 0.91 | 1.04 |
| Weight of the edit in the slope | | | 0.00 | 0.03 | 0.05 | 0.01 | 0.72 | 0.20 |

Mean system change regressed through the origin on the mean reference change across the six clinical edits (log-odds scale; mean over supported evaluation records); 1 means the system moves as much as the reference. Simultaneous 95% intervals across 66 tests (60 edit-by-system cells, from these six edits and four demographic edits not shown, and 6 clinical response slopes; critical value 3.23, including the factor 1.05). ‘Without’ columns: slope (spline reference) with that edit left out. Weights: share of the slope’s information from each edit, the same for every system.

### Supplementary Table 11. Repeatability and prior exposure

#### a. Three calls with identical input

| System | Risk: ICC (95% CI) | Risk: identical, % | Patient questions: Fleiss' kappa (95% CI) | Patient questions: identical, % | Staging: ICC (95% CI) | Staging: identical, % | Kidney injury: ICC (95% CI) | Kidney injury: identical, % |
|---|---|---|---|---|---|---|---|---|
| Jev 1.13 | 0.996 (0.995–0.997) | 16 | 0.97 (0.90–1.00) | 98 | 0.990 (0.981–0.997) | 82 | 1.000 | 100 |
| GPT-5.6 Sol | 0.992 (0.990–0.993) | 3 | 1.00 | 100 | 1.000 | 100 | 0.975 (0.932–1.000) | 92 |
| Claude Opus 5.5 | 0.996 (0.995–0.997) | 27 | 1.00 | 100 | 1.000 | 100 | 1.000 | 100 |
| Gemini 3.8 Flash | 1.000 (0.999–1.000) | 92 | 1.00 | 100 | 1.000 | 100 | 1.000 | 100 |
| Muse Spark 1.1 | 0.988 (0.985–0.990) | 1 | 0.96 (0.88–1.00) | 98 | 1.000 | 100 | 0.955 (0.849–1.000) | 95 |
| GLM-5.3 | 0.973 (0.966–0.978) | 0 | 1.00 | 100 | 0.996 (0.990–1.000) | 92 | 1.000 | 100 |
| GPT-5.6 Luna | — | — | 1.00 | 100 | 0.998 (0.993–1.000) | 95 | 0.945 (0.851–0.993) | 90 |
| Claude Sonnet 5 | — | — | 0.88 (0.74–1.00) | 92 | 0.999 (0.996–1.000) | 98 | 0.969 (0.926–0.994) | 90 |
| Gemini 3.5 Flash-Lite | — | — | 1.00 | 100 | 1.000 | 100 | 1.000 | 100 |
| MedGemma 27B | — | — | 0.97 (0.90–1.00) | 98 | 0.805 (0.637–0.910) | 55 | 0.849 (0.729–0.931) | 70 |
| Gemma 3 27B | — | — | 0.93 (0.83–1.00) | 95 | 0.913 (0.836–0.962) | 62 | 0.969 (0.911–1.000) | 95 |

Ten-year risk: 358 baseline and edited records of 100 participants, log-odds scale; patient questions: 40; staging: 40 reports; kidney injury: 40 stays; level index for the last two. ICC: two-way random effects, absolute agreement, single call. Jev in the recommended form for cancer staging gave the same stage on every repeat of 40 reports, in both versions. The smaller LLMs answered the ten-year risk records once, so they have no risk repeatability.

#### b. Masked-field recovery

| Data | System | Records | Exact recovery, % | Ratio to the imputation model | Possible exposure |
|---|---|---|---|---|---|
| NHANES | GPT-5.6 Sol | 200 | 0.5 | 0.50 | no |
| NHANES | Claude Opus 5.5 | 200 | 2.5 | 2.50 | no |
| NHANES | Gemini 3.8 Flash | 200 | 0.5 | 0.50 | no |
| NHANES | Muse Spark 1.1 | 200 | 1.5 | 1.50 | no |
| NHANES | GLM-5.3 | 200 | 1.0 | 1.00 | no |
| NHANES | Imputation model | 200 | 1.0 | 1.00 | no |
| eICU demo | GPT-5.6 Sol | 200 | 3.0 | 0.86 | no |
| eICU demo | Claude Opus 5.5 | 200 | 4.0 | 1.14 | no |
| eICU demo | Gemini 3.8 Flash | 200 | 4.5 | 1.29 | no |
| eICU demo | Muse Spark 1.1 | 200 | 4.5 | 1.29 | no |
| eICU demo | GLM-5.3 | 200 | 2.5 | 0.71 | no |
| eICU demo | Imputation model | 200 | 3.5 | 1.00 | no |

One field was masked in each of 200 records (40 records for each of five fields: in NHANES mean cell volume, alkaline phosphatase, total cholesterol, lymphocyte percentage and waist circumference; in eICU heart rate, sodium, haematocrit, urea nitrogen and creatinine) and the system was asked for its value. Possible exposure: exact recovery of at least 5% and more than twice the imputation model's. Jev was not asked, having no free-text answer.

### Supplementary Table 12. Risk of death: the named categories of the recommended form

#### a. Ten-year risk records

| Field in the record | Sent as | Categories (cut points) |
|---|---|---|
| age | age group | 40 to 44 years (< 45); 45 to 49 years (< 50); 50 to 54 years (< 55); 55 to 59 years (< 60); 60 to 64 years (< 65); 65 to 69 years (< 70); 70 to 74 years (< 75); 75 to 79 years (≥ 75) [years] |
| body mass index | body mass index category | underweight (< 18.5); normal weight (< 25); pre-obesity (< 30); obesity class I (< 35); obesity class II (< 40); obesity class III (≥ 40) [kg/m²] |
| waist circumference | waist circumference category | men: no abdominal obesity (≤ 102); abdominal obesity (> 102) [cm]; women: no abdominal obesity (≤ 88); abdominal obesity (> 88) [cm] |
| systolic blood pressure, diastolic blood pressure | blood pressure category | stage 2 hypertension (systolic ≥ 140 or diastolic ≥ 90); stage 1 hypertension (systolic ≥ 130 or diastolic ≥ 80); elevated blood pressure (systolic ≥ 120); normal blood pressure (otherwise) [mmHg]; the higher category when the two readings differ |
| HbA1c | HbA1c category | below the prediabetes range (< 5.7); prediabetes range (< 6.5); diabetes range (≥ 6.5) [%] |
| serum glucose random | serum glucose random category | below the ADA random-glucose threshold for diabetes (< 200); at or above the ADA random-glucose threshold for diabetes (≥ 200) [mg/dL] |
| serum creatinine | estimated glomerular filtration rate category | KDIGO G5, kidney failure (< 15); KDIGO G4, severely decreased (< 30); KDIGO G3b, moderately to severely decreased (< 45); KDIGO G3a, mildly to moderately decreased (< 60); KDIGO G2, mildly decreased (< 90); KDIGO G1, normal or high (≥ 90) [mL/min/1.73 m²]; eGFR computed in code from creatinine, age and sex (CKD-EPI 2021) |
| urine albumin-to-creatinine ratio | urine albumin-to-creatinine ratio category | KDIGO A1, normal to mildly increased (< 30); KDIGO A2, moderately increased (≤ 300); KDIGO A3, severely increased (> 300) [mg/g] |
| serum albumin | serum albumin category | below the reference range (< 3.5); within the reference range (≤ 5.5); above the reference range (> 5.5) [g/dL] |
| white blood cell count | white blood cell count category | below the reference range (< 4); within the reference range (≤ 11); above the reference range (> 11) [×10³/µL] |
| lymphocyte percent | lymphocyte percent category | below the reference range (< 30); within the reference range (≤ 45); above the reference range (> 45) [%] |
| mean corpuscular volume | mean corpuscular volume category | below the reference range (< 80); within the reference range (≤ 98); above the reference range (> 98) [fL] |
| red cell distribution width | red cell distribution width category | below the reference range (< 9); within the reference range (≤ 14.5); above the reference range (> 14.5) [%] |
| alkaline phosphatase | alkaline phosphatase category | below the reference range (< 30); within the reference range (≤ 120); above the reference range (> 120) [U/L] |
| C-reactive protein | C-reactive protein category | low relative cardiovascular risk (< 1); average relative cardiovascular risk (≤ 3); high relative cardiovascular risk (≤ 10); markedly raised, suggesting infection or inflammation (> 10) [mg/L]; the record's value ×10 (mg/dL to mg/L) |
| total cholesterol | total cholesterol category | desirable (< 200); borderline high (< 240); high (≥ 240) [mg/dL] |
| HDL cholesterol | HDL cholesterol category | low (< 40); neither low nor high (< 60); high (≥ 60) [mg/dL] |
| prescription medicines past 30 days | prescription medicines past 30 days category | none (< 1); 1 to 4 (< 5); 5 or more (polypharmacy) (≥ 5) [medicines] |
| family income-to-poverty ratio | family income-to-poverty ratio category | below 100% of the poverty threshold (< 1); 100% to 199% of the poverty threshold (< 2); 200% to 399% of the poverty threshold (< 4); 400% or more of the poverty threshold (≥ 4) [× poverty threshold] |

Fields sent unchanged: record type, source, sex, race and ethnicity, smoking status, self-rated health, doctor-diagnosed conditions, education, has health insurance. A missing field stayed missing. Each value was placed in a category as the original record showed it; the category's name, never the value or its cut points, was sent. Sources: WHO adult body-mass-index classes[3]; NCEP ATP III (waist, total and HDL cholesterol)[4]; 2017 ACC/AHA blood-pressure guideline[5]; ADA Standards of Care 2024 (HbA1c, random glucose)[6]; CKD-EPI 2021 equation[7] with KDIGO 2012 GFR and albuminuria categories[8]; ABIM laboratory reference ranges (January 2026)[9]; AHA/CDC statement on C-reactive protein[10]; polypharmacy as five or more medicines; NCHS income-to-poverty categories.

**b. Intensive care records**

| Field | Categories sent | Source |
|---|---|---|
| Age | 18 to 19 years … 85 years and over | NHANES source, unchanged |
| Serum creatinine | KDIGO G1 to G5 (eGFR by CKD-EPI 2021) | NHANES source, unchanged |
| Serum glucose | As in part a | NHANES source, unchanged |
| Serum albumin | As in part a | NHANES source, unchanged |
| White blood cell count | As in part a | NHANES source, unchanged |
| Serum sodium | below the reference range (<136); within the reference range (≤145); above the reference range | ABIM laboratory reference ranges, January 2026 |
| Haematocrit | female: below the reference range (<37), within the reference range (≤47), above the reference range; male: below the reference range (<42), within the reference range (≤50), above the reference range | ABIM laboratory reference ranges, January 2026 |
| Blood urea nitrogen | below the reference range (<8); within the reference range (≤20); above the reference range | ABIM laboratory reference ranges, January 2026 |
| Serum bilirubin | below the reference range (<0.3); within the reference range (≤1); above the reference range | ABIM laboratory reference ranges, January 2026 |
| Arterial pH | below the reference range (<7.38); within the reference range (≤7.44); above the reference range | ABIM laboratory reference ranges, January 2026 |
| Arterial PO2, inspired oxygen fraction | SOFA respiration score 4 (<100) with ventilation; SOFA respiration score 3 (<200) with ventilation; SOFA respiration score 2 (<300); SOFA respiration score 1 (<400); SOFA respiration score 0 | Sepsis-3 consensus (SOFA, SIRS, lactate) |
| Mean arterial pressure | SOFA cardiovascular score 1 by mean arterial pressure (<70); SOFA cardiovascular score 0 by mean arterial pressure | Sepsis-3 consensus (SOFA, SIRS, lactate) |
| Glasgow coma scale | SOFA central nervous system score 4 (<6); SOFA central nervous system score 3 (<10); SOFA central nervous system score 2 (<13); SOFA central nervous system score 1 (<15); SOFA central nervous system score 0 | Sepsis-3 consensus (SOFA, SIRS, lactate) |
| Urine output 24 h | SOFA renal score 4 by urine output (<200); SOFA renal score 3 by urine output (<500); above the SOFA urine-output thresholds | Sepsis-3 consensus (SOFA, SIRS, lactate) |
| Temperature | below the SIRS temperature range (<36); within the SIRS temperature range (≤38); above the SIRS temperature range | Sepsis-3 consensus (SOFA, SIRS, lactate) |
| Heart rate | at or below the SIRS heart-rate threshold (≤90); above the SIRS heart-rate threshold | Sepsis-3 consensus (SOFA, SIRS, lactate) |
| Respiratory rate | at or below the SIRS respiratory-rate threshold (≤20); above the SIRS respiratory-rate threshold | Sepsis-3 consensus (SOFA, SIRS, lactate) |
| Arterial PCO2 | below the SIRS PaCO2 threshold (<32); at or above the SIRS PaCO2 threshold | Sepsis-3 consensus (SOFA, SIRS, lactate) |
| Maximum lactate | at or below the Sepsis-3 lactate threshold (≤2); above the Sepsis-3 lactate threshold | Sepsis-3 consensus (SOFA, SIRS, lactate) |

Each numeric field was replaced by the name of its category; other fields were sent unchanged and a missing value stayed missing. Rule: a field the NHANES records also have keeps its NHANES source; other laboratory values use the NHANES laboratory source; fields recorded only in intensive care use the Sepsis-3 consensus[11], and arterial pH, which it does not categorise, the laboratory source.

### Supplementary Table 13. Clinical decisions: sensitivity, specificity and AUROC

#### a. Patient questions: ordering the test or treatment

Ordering the test or treatment (55 indicated, 125 low-value questions): sensitivity, ordering when indicated; specificity, holding off when low-value.

| System | Sensitivity, % (95% CI) | Specificity, % (95% CI) | AUROC |
|---|---|---|---|
| Jev 1.13 | 94.5 (85.1–98.1) | 80.8 (73.0–86.7) | 0.962 |
| GPT-5.6 Sol | 100.0 (93.5–100.0) | 100.0 (97.0–100.0) | 1.000 |
| Claude Opus 5.5 | 100.0 (93.5–100.0) | 100.0 (97.0–100.0) | 1.000 |
| Gemini 3.8 Flash | 100.0 (93.5–100.0) | 99.2 (95.6–99.9) | 0.999 |
| Muse Spark 1.1 | 98.2 (90.4–99.7) | 100.0 (97.0–100.0) | 1.000 |
| GLM-5.3 | 100.0 (93.5–100.0) | 96.8 (92.1–98.7) | 0.998 |
| GPT-5.6 Luna | 100.0 (93.5–100.0) | 99.2 (95.6–99.9) | 1.000 |
| Claude Sonnet 5 | 98.2 (90.4–99.7) | 96.8 (92.1–98.7) | 0.991 |
| Gemini 3.5 Flash-Lite | 98.2 (90.4–99.7) | 92.8 (86.9–96.2) | 0.966 |
| MedGemma 27B | 94.5 (85.1–98.1) | 77.6 (69.5–84.0) | 0.877 |
| Gemma 3 27B | 96.4 (87.7–99.0) | 73.6 (65.3–80.5) | 0.850 |

#### b. Cancer staging

Stage III or IV, against stage 0–II (124 positive, 116 negative reports of the 240):

| System | Stage names only: sensitivity, % (95% CI) | Specificity, % (95% CI) | AUROC | Medical context: sensitivity, % (95% CI) | Specificity, % (95% CI) | AUROC |
|---|---|---|---|---|---|---|
| Jev 1.13 | 92.7 (86.8–96.1) | 100.0 (96.8–100.0) | 0.999 | 97.6 (93.1–99.2) | 100.0 (96.8–100.0) | 1.000 |
| Jev 1.13, recommended form | 100.0 (97.0–100.0) | 100.0 (96.8–100.0) | 1.000 | 100.0 (97.0–100.0) | 100.0 (96.8–100.0) | 1.000 |
| GPT-5.6 Sol | 100.0 (97.0–100.0) | 100.0 (96.8–100.0) | 1.000 | 100.0 (97.0–100.0) | 100.0 (96.8–100.0) | 1.000 |
| Claude Opus 5.5 | 100.0 (97.0–100.0) | 100.0 (96.8–100.0) | 1.000 | 100.0 (97.0–100.0) | 100.0 (96.8–100.0) | 1.000 |
| Gemini 3.8 Flash | 100.0 (97.0–100.0) | 100.0 (96.8–100.0) | 1.000 | 100.0 (97.0–100.0) | 100.0 (96.8–100.0) | 1.000 |
| Muse Spark 1.1 | 100.0 (97.0–100.0) | 100.0 (96.8–100.0) | 1.000 | 100.0 (97.0–100.0) | 100.0 (96.8–100.0) | 1.000 |
| GLM-5.3 | 92.7 (86.8–96.1) | 98.3 (93.9–99.5) | 1.000 | 96.0 (90.9–98.3) | 99.1 (95.3–99.8) | 1.000 |
| GPT-5.6 Luna | 100.0 (97.0–100.0) | 100.0 (96.8–100.0) | 1.000 | 100.0 (97.0–100.0) | 100.0 (96.8–100.0) | 1.000 |
| Claude Sonnet 5 | 100.0 (97.0–100.0) | 100.0 (96.8–100.0) | 1.000 | 100.0 (97.0–100.0) | 100.0 (96.8–100.0) | 1.000 |
| Gemini 3.5 Flash-Lite | 99.2 (95.6–99.9) | 96.6 (91.5–98.7) | 0.983 | 98.4 (94.3–99.6) | 100.0 (96.8–100.0) | 0.996 |
| MedGemma 27B | 92.7 (86.8–96.1) | 55.2 (46.1–63.9) | 0.764 | 100.0 (97.0–100.0) | 44.0 (35.3–53.0) | 0.717 |
| Gemma 3 27B | 95.2 (89.8–97.8) | 45.7 (36.9–54.7) | 0.807 | 100.0 (97.0–100.0) | 32.8 (24.9–41.7) | 0.855 |

Stage IV, against stage 0–III (61 positive, 179 negative reports):

| System | Stage names only: sensitivity, % (95% CI) | Specificity, % (95% CI) | AUROC | Medical context: sensitivity, % (95% CI) | Specificity, % (95% CI) | AUROC |
|---|---|---|---|---|---|---|
| Jev 1.13 | 70.5 (58.1–80.4) | 100.0 (97.9–100.0) | 0.999 | 63.9 (51.4–74.8) | 100.0 (97.9–100.0) | 1.000 |
| Jev 1.13, recommended form | 44.3 (32.5–56.7) | 100.0 (97.9–100.0) | 0.958 | 44.3 (32.5–56.7) | 100.0 (97.9–100.0) | 0.958 |
| GPT-5.6 Sol | 100.0 (94.1–100.0) | 100.0 (97.9–100.0) | 1.000 | 100.0 (94.1–100.0) | 100.0 (97.9–100.0) | 1.000 |
| Claude Opus 5.5 | 100.0 (94.1–100.0) | 100.0 (97.9–100.0) | 1.000 | 100.0 (94.1–100.0) | 100.0 (97.9–100.0) | 1.000 |
| Gemini 3.8 Flash | 100.0 (94.1–100.0) | 100.0 (97.9–100.0) | 1.000 | 100.0 (94.1–100.0) | 100.0 (97.9–100.0) | 1.000 |
| Muse Spark 1.1 | 100.0 (94.1–100.0) | 100.0 (97.9–100.0) | 1.000 | 100.0 (94.1–100.0) | 100.0 (97.9–100.0) | 1.000 |
| GLM-5.3 | 98.4 (91.3–99.7) | 94.4 (90.0–96.9) | 1.000 | 98.4 (91.3–99.7) | 97.2 (93.6–98.8) | 1.000 |
| GPT-5.6 Luna | 100.0 (94.1–100.0) | 100.0 (97.9–100.0) | 1.000 | 100.0 (94.1–100.0) | 100.0 (97.9–100.0) | 1.000 |
| Claude Sonnet 5 | 100.0 (94.1–100.0) | 100.0 (97.9–100.0) | 1.000 | 100.0 (94.1–100.0) | 100.0 (97.9–100.0) | 1.000 |
| Gemini 3.5 Flash-Lite | 96.7 (88.8–99.1) | 100.0 (97.9–100.0) | 0.983 | 95.1 (86.5–98.3) | 99.4 (96.9–99.9) | 0.973 |
| MedGemma 27B | 91.8 (82.2–96.4) | 69.8 (62.7–76.1) | 0.844 | 68.9 (56.4–79.1) | 92.7 (88.0–95.7) | 0.823 |
| Gemma 3 27B | 77.0 (65.1–85.8) | 90.5 (85.3–94.0) | 0.923 | 32.8 (22.3–45.3) | 100.0 (97.9–100.0) | 0.832 |

Jev in the recommended form answered the same four questions whichever version the LLMs saw; its row repeats in both column groups. An unparsable answer counts as wrong. AUROC from the probability summed over the positive group's levels; for the four-question form, over the stage groups computed from the four answers' probabilities.

#### c. Kidney-injury stage 2 or 3

Stage 2 or 3, against no injury or stage 1 (83 positive, 478 negative stays):

| System | Names only: sensitivity, % (95% CI) | Specificity, % (95% CI) | AUROC | KDIGO criteria: sensitivity, % (95% CI) | Specificity, % (95% CI) | AUROC |
|---|---|---|---|---|---|---|
| Jev 1.13 | 94.0 (86.7–97.4) | 70.5 (66.3–74.4) | 0.918 | 95.2 (88.3–98.1) | 97.1 (95.1–98.2) | 0.990 |
| GPT-5.6 Sol | 98.8 (93.5–99.8) | 97.5 (95.7–98.6) | 0.995 | 100.0 (95.6–100.0) | 100.0 (99.2–100.0) | 1.000 |
| Claude Opus 5.5 | 97.6 (91.6–99.3) | 100.0 (99.2–100.0) | 0.999 | 98.8 (93.5–99.8) | 100.0 (99.2–100.0) | 0.997 |
| Gemini 3.8 Flash | 98.8 (93.5–99.8) | 97.9 (96.2–98.9) | 0.998 | 100.0 (95.6–100.0) | 100.0 (99.2–100.0) | 1.000 |
| Muse Spark 1.1 | 96.4 (89.9–98.8) | 99.4 (98.2–99.8) | 0.999 | 100.0 (95.6–100.0) | 100.0 (99.2–100.0) | 1.000 |
| GLM-5.3 | 88.0 (79.2–93.3) | 91.6 (88.8–93.8) | 0.999 | 97.6 (91.6–99.3) | 97.9 (96.2–98.9) | 1.000 |
| GPT-5.6 Luna | 96.4 (89.9–98.8) | 96.0 (93.9–97.4) | 0.977 | 94.0 (86.7–97.4) | 99.0 (97.6–99.6) | 0.981 |
| Claude Sonnet 5 | 91.6 (83.6–95.9) | 98.1 (96.5–99.0) | 0.962 | 92.8 (85.1–96.6) | 99.6 (98.5–99.9) | 0.988 |
| Gemini 3.5 Flash-Lite | 83.1 (73.7–89.7) | 86.2 (82.8–89.0) | 0.906 | 80.7 (71.0–87.8) | 94.1 (91.7–95.9) | 0.917 |
| MedGemma 27B | 80.7 (71.0–87.8) | 85.8 (82.4–88.6) | 0.870 | 83.1 (73.7–89.7) | 87.7 (84.4–90.3) | 0.888 |
| Gemma 3 27B | 81.9 (72.3–88.7) | 83.9 (80.3–86.9) | 0.864 | 81.9 (72.3–88.7) | 85.6 (82.1–88.4) | 0.871 |

### Supplementary Note 1. Prompts and reading of replies

Within each task, Jev and every LLM received the same case text, question and answer options. LLMs also received the reply format below and field labels separating the case from the question. Braces mark the parts filled per item.

#### Ten-year risk of death

**LLMs**, system message:

> You are helping with a research study of mortality risk. You will see one de-identified record from a national health survey and one statement about the person it describes. Give the probability that the statement is true, as a number between 0 and 1 with three decimal places. Always give a number. Reply with a JSON object whose only key is "p", and nothing else.

User message:

> Record:
> {record}
>
> Statement: This person will die from any cause within ten years of this examination.

Reply schema, where the provider supports one: an object with a single number p between 0 and 1.

**Jev**: a Noul question with the record as the case and the same statement; Jev returns the probability that the statement is true.

Recommended form, intensive care records (Jev only; every evaluation stay): the statement "This patient will die before hospital discharge." unchanged, with each numeric field of the record replaced by the name of its category (Supplementary Table 12b).

Masked-field test (LLMs only; results in Supplementary Table 11), system message:

> You are helping with a research study. In the de-identified record below, one field's value has been replaced by "MISSING". Give the value you think this record most likely contains, in the unit shown. Always give a number. Reply with a JSON object whose only key is "value", and nothing else.

#### Patient questions

**LLMs**, system message:

> Choose the option that is the most appropriate next step, and give the probability that each option is the most appropriate next step, as numbers between 0 and 1 that add up to 1. Always choose one option. Reply with a JSON object with two keys, "choice" (the letter of the option you choose) and "probabilities" (an object with one number for each letter), and nothing else.

User message:

> Message:
> {patient question}
>
> Question: Which is the most appropriate next step for this person?
>
> A. {option}
> B. {option}
> C. {option}
> D. {option}

Reply schema: choice (one of A–D) and probabilities (one number between 0 and 1 for each letter).

Example user message (a low-value question under recommendation 1 of Supplementary Table 2c; the options in the order shown for this question; key D):

> Message:
> I'm 34, male. About two weeks ago I started getting pain in my lower back after helping my friend move apartments. The pain is mostly on the right side and gets worse when I sit for a long time at work. I can still walk and do most things, but it's really uncomfortable. I've been taking ibuprofen which helps a little. I'm worried this could be a herniated disc or something serious. What should I do? Do I need any tests or should I see a specialist?
>
> Question: Which is the most appropriate next step for this person?
>
> A. Start an opioid painkiller and review in two weeks.
> B. Arrange imaging of the lumbar spine (MRI or CT) now.
> C. Start a short course of oral steroids and review in two weeks.
> D. No imaging for now: stay active, use simple pain relief, and review if the pain or symptoms change.

**Jev**: a Choice question with the patient question as the case, the same question and the four options, keyed by the same letters in the same order.

**Board questions**

**LLMs**, system message:

> Answer this medical question, and give the probability that each option is the correct answer, as numbers between 0 and 1 that add up to 1. Always choose one option. Reply with a JSON object with two keys, “choice” (the letter of the option you choose) and “probabilities” (an object with one number for each letter), and nothing else.

User message:

> {question}
>
> A. {option}
> B. {option}
> …

One line per option, four or five as published. Reply schema as for the patient questions, with one number for each letter.

**Jev**: a Choice question with the question as the case, the instruction “Answer this medical question.” and the options keyed by the same letters in the same order.

**Cancer staging**

**LLMs**, system message:

> Choose the stage group that is correct, and give the probability that each stage group is correct, as numbers between 0 and 1 that add up to 1. Always choose one stage group. Reply with a JSON object with two keys, “stage” (the stage group you choose, written exactly as in the list) and “probabilities” (an object with one number for each stage group), and nothing else.

User message:

> Pathology report:
> {report}
>
> Question: Assign the pathological stage group for this colorectal cancer according to the AJCC Cancer Staging Manual, 8th edition. Stage IVA-IVB here includes IVA and IVB.
>
> Stage groups:
> {the ten levels, one per line}

Example report (ordinary, synoptic layout; key IIIB: T3 N1a M0, 1 of 16 regional nodes involved):

> SURGICAL PATHOLOGY REPORT
> Specimen: Ascending colon, right hemicolectomy
>
> CLINICAL INFORMATION
> Tumor in the ascending colon seen at colonoscopy, referred for resection.
> Staging CT chest, abdomen and pelvis: Negative for distant metastases.
>
> COLON RESECTION: SYNOPTIC REPORT
> Procedure: Laparoscopic right hemicolectomy
> Tumor site: Ascending colon
> Histologic type: Adenocarcinoma, not otherwise specified
> Histologic grade: Low grade (moderately differentiated)
> Tumor size: 2.8 x 2.1 x 1.7 cm
> Tumor extent: Beyond the muscularis propria into subserosal adipose tissue, with the overlying visceral peritoneum intact
> Macroscopic tumor perforation: Absent
> Lymphovascular invasion: Present (small vessel)
> Perineural invasion: Not identified
> Margins: All margins negative, closest margin at 2.7 cm
> Regional lymph nodes: 1 of 16 involved
> Tumor deposits: Not identified
> Mismatch repair immunohistochemistry: Retained nuclear expression of MLH1, PMS2, MSH2 and MSH6

In the version with medical context, each stage is followed by its definition (Supplementary Table 3a), for example “IIIB: T3-T4a N1a-N1c M0; T2-T3 N2a M0; T1-T2 N2b M0”. Reply schema: stage (one of the ten levels) and probabilities (one number for each level).

**Jev**: a Score question with the report as the case, the same question and the ten levels, lowest first, as the ordered scale.

**Four-question forms, LLMs** (task decomposition only; both, without examples; the recommended form; the recommended form with AJCC notes), system message:

> For each question, choose the option that is correct, and give the probability that each option is correct, as numbers between 0 and 1 that add up to 1. Always choose one option for each question. Reply with a JSON object with one key for each question ID, each holding an object with two keys, “choice” (the option you choose, written exactly as in the question’s criteria) and “probabilities” (an object with one number for each option), and nothing else.

User message:

> Pathology report:
> {report}
>
> Questions:
> {Jev's four questions, exactly as sent to Jev, as JSON}

The questions object (question IDs, instructions, options in order, and each option’s definition, exclusions and examples where the form has them) is the one Jev received for that form (Supplementary Note 2). Reply schema: one object per question ID, each with choice (one of that question’s options) and probabilities (one number for each option). The stage group was computed in code from the four choices, and confidence was the product of the four top probabilities, as for Jev.

**Jev**: the same four Choice questions, with the report as a named field (pathology_report).

#### Kidney-injury staging

**LLMs**, system message:

> Choose the AKI stage that is correct, and give the probability that each AKI stage is correct, as numbers between 0 and 1 that add up to 1. Always choose one AKI stage. Reply with a JSON object with two keys, “stage” (the AKI stage you choose, written exactly as in the list) and “probabilities” (an object with one number for each AKI stage), and nothing else.

User message (an example record):

> Patient:
> Age: 57 years
> Sex: female
> Admission weight: 158.8 kg
> Serum creatinine (mg/dL), from 7 days before to 72 hours after intensive care unit admission; times are from unit admission:
> -3 h 25 min: 1.62
> +15 h 40 min: 1.56
> +40 h 09 min: 1.97
> +63 h 34 min: 1.45
> Dialysis started: no
>
> Question: By the KDIGO creatinine criteria, what was this patient's highest AKI stage during the first 72 hours in the intensive care unit?
>
> AKI stages:
> no AKI
> stage 1
> stage 2
> stage 3

In the version with KDIGO criteria, each stage is followed by its criteria: no AKI, “no value from unit admission to 72 hours is 0.3 mg/dL or more above a value in the preceding 48 hours or 1.5 times or more the lowest value in the preceding 7 days, and no dialysis started”; stage 1, “a value 1.5 times or more but less than 2.0 times the lowest value in the preceding 7 days, or 0.3 mg/dL or more above a value in the preceding 48 hours”; stage 2, “a value 2.0 times or more but less than 3.0 times the lowest value in the preceding 7 days”; stage 3, “a value 3.0 times or more the lowest value in the preceding 7 days; or a value of 4.0 mg/dL or more that itself meets the stage 1 rise or ratio; or dialysis started”. Reply schema: stage (one of the four) and probabilities (one number for each).

**Jev**: a Score question with the record as the case, the same question and the four stages, lowest first, as the ordered scale.

#### Reading replies

One rule applied to every system. A reply had to be one JSON object (a code fence was allowed), except in the board questions, where a reply that was not one JSON object was read from the last JSON object in it holding a choice and probabilities; only GLM-5.3 gave such replies (285 of its 3,142 answers). A label matched exactly, then ignoring case, then after dropping a leading word such as “option” or “stage”, brackets or a trailing full stop. In cancer staging, an answer of IVA or IVB counted as IVA–IVB. A level or option missing from the probabilities counted as zero. Probability lists summing to between 0.9 and 1.1 were rescaled to sum to one; other lists were left out of the confidence analysis only. A reply with no readable answer counted as missing in ten-year risk and as wrong in every other task.

#### Style check (patient questions)

Word count: tokens separated by spaces. Negations: case-insensitive matches of \b(never|no|not|none|nothing|without|cannot|nobody|nowhere|neither|nor|dont|cant|wont|didnt|doesnt|isnt|wasnt|\w+n['’]t)\b.

### Supplementary Note 2. Recommended form for cancer staging: the questions and options as sent

The report was sent whole as the one field of the case (pathology_report). The four questions below went in one request; only the report changed between requests. Text in *italics* was sent only in the second version (recommended form with AJCC notes), appended to the option's definition or exclusion. The stage group was not asked: it was computed in code from the four answers with the AJCC 8th edition table that built the key, involved nodes being asked in the bands that define N. Jev's confidence was the product of the four top probabilities. The format follows the developer's documentation (docs.typesafe.ai: case as a JSON object with named fields; every question in one request; options defined by what they are, what they are not for and examples; numbers in named bands; arithmetic in code).

#### T category

Question: "Which pathological T category does this colorectal cancer have, according to the AJCC Cancer Staging Manual, 8th edition?"

| Option | What it is | Not for | Examples |
|---|---|---|---|
| Tis | Carcinoma in situ or intramucosal carcinoma: tumor involves the lamina propria, with no extension through the muscularis mucosae. | Tumor that extends through the muscularis mucosae into the submucosa or deeper. | "Intramucosal adenocarcinoma; no invasion of the submucosa." / "Tumor extent: lamina propria (intramucosal)" |
| T1 | Tumor invades the submucosa, through the muscularis mucosae but not into the muscularis propria. | Tumor that invades the muscularis propria or deeper. | "Adenocarcinoma invades the submucosa; the muscularis propria is not involved." / "Tumor extent: submucosa" |
| T2 | Tumor invades the muscularis propria, without extending through it. | Tumor that extends through the muscularis propria into the pericolorectal tissues. | "Invasive carcinoma is confined to the muscularis propria." / "Tumor extent: muscularis propria" |
| T3 | Tumor invades through the muscularis propria into the pericolorectal tissues, without reaching the surface of the visceral peritoneum and without invading another organ or structure. | Tumor at the surface of the visceral peritoneum; macroscopic perforation of the bowel through the tumor; tumor that invades another organ or structure. | "Adenocarcinoma penetrates the muscularis propria and infiltrates the pericolorectal fat." / "Tumor extent: through the muscularis propria into pericolic fat" |
| T4a | Tumor invades through the visceral peritoneum to its surface, including macroscopic perforation of the bowel through the tumor and continuous invasion of tumor through areas of inflammation to the surface of the visceral peritoneum. | Tumor that invades another organ or structure. | "Tumor cells are present at the serosal (visceral peritoneal) surface." / "Tumor extent: reaches the visceral peritoneal surface" |
| T4b | Tumor directly invades another organ or structure. *Tumor that is adherent to another organ or structure counts as T4b only when tumor is present in the adhesion on microscopy.* | Tumor limited to the bowel wall and the visceral peritoneum. *An adhesion to another organ or structure with no tumor in it on microscopy, which is classified by the depth of invasion.* | "Carcinoma directly invades the wall of the urinary bladder." / "Tumor extent: invades the adjacent small bowel" |
| cannot be assessed | The report does not allow the depth of invasion of the primary tumor to be assessed. | A report that states how deep the primary tumor invades. | "Depth of invasion cannot be determined (fragmented specimen)." |

#### Regional nodes

Question: “How many regional lymph nodes contain metastatic carcinoma (tumor in the node measuring 0.2 mm or more), according to the AJCC Cancer Staging Manual, 8th edition?”

| Option | What it is | Not for | Examples |
|---|---|---|---|
| none | No regional lymph node contains metastatic carcinoma. *Nodes with isolated tumor cells only (single cells or clusters smaller than 0.2 mm) count as negative.* | A report in which at least one regional lymph node contains metastatic carcinoma. | “No metastatic carcinoma in any of twenty-two regional lymph nodes (0/22).” / “Lymph nodes: 0 positive / 18 examined” |
| 1 | Exactly one regional lymph node contains metastatic carcinoma. *Nodes with isolated tumor cells only and non-regional lymph nodes (for example, para-aortic, aortocaval or other retroperitoneal nodes) are not counted; a metastasis in a non-regional lymph node is distant metastasis.* | No involved regional node, or two or more involved regional nodes. | “One of nineteen regional lymph nodes contains metastatic adenocarcinoma (1/19).” / “Lymph nodes: 1 positive / 14 examined” |
| 2 to 3 | Two or three regional lymph nodes contain metastatic carcinoma. *Nodes with isolated tumor cells only and non-regional lymph nodes (for example, para-aortic, aortocaval or other retroperitoneal nodes) are not counted; a metastasis in a non-regional lymph node is distant metastasis.* | Fewer than two or more than three involved regional nodes. | “Metastatic carcinoma in three of twenty-one regional lymph nodes (3/21).” / “Lymph nodes: 2 positive / 16 examined” |
| 4 to 6 | Four, five or six regional lymph nodes contain metastatic carcinoma. *Nodes with isolated tumor cells only and non-regional lymph nodes (for example, para-aortic, aortocaval or other retroperitoneal nodes) are not counted; a metastasis in a non-regional lymph node is distant metastasis.* | Fewer than four or more than six involved regional nodes. | “Five of eighteen regional lymph nodes contain metastatic adenocarcinoma (5/18).” / “Lymph nodes: 4 positive / 23 examined” |
| 7 or more | Seven or more regional lymph nodes contain metastatic carcinoma. *Nodes with isolated tumor cells only and non-regional lymph nodes (for example, para-aortic, aortocaval or other retroperitoneal nodes) are not counted; a metastasis in a non-regional lymph node is distant metastasis.* | Six or fewer involved regional nodes. | “Metastatic carcinoma in nine of twenty-four regional lymph nodes (9/24).” / “Lymph nodes: 11 positive / 20 examined” |
| not stated | The report does not give the number of regional lymph nodes that contain metastatic carcinoma. | A report that gives the number of involved regional nodes, including zero. | “Lymph nodes: see separate report” |

#### Tumor deposits

Question: “Does the report describe tumor deposits, according to the AJCC Cancer Staging Manual, 8th edition?”

| Option | What it is | Not for | Examples |
|---|---|---|---|
| present | The report records one or more tumor deposits: discrete tumor nodules in the pericolorectal fat or mesentery, separate from the primary tumor, without identifiable lymph node tissue or identifiable vascular or neural structure. | A report that states tumor deposits are absent or does not mention them. | “Two discrete tumor nodules in the mesenteric fat, without residual lymph node tissue (tumor deposits).” / “Tumor deposits: present (number: 2)” |
| absent | The report states that no tumor deposits are present. | A report that describes one or more tumor deposits. | “Tumor deposits: none seen” |
| not stated | The report neither mentions tumor deposits nor describes such nodules. | A report that states whether tumor deposits are present. | “A report whose lymph node section gives the node count and says nothing about tumor deposits.” |

**Distant metastasis**

Question: “Which distant metastasis category do the pathology findings and any staging imaging in the report describe, according to the AJCC Cancer Staging Manual, 8th edition?”

| Option | What it is | Not for | Examples |
|---|---|---|---|
| none | No distant metastasis. | Metastasis in one or more distant sites or organs, or in the peritoneum. *A metastasis in a non-regional lymph node (for example, para-aortic, aortocaval or other retroperitoneal nodes), which is distant metastasis in one site (M1a).* | “CT chest, abdomen and pelvis: no metastatic disease.” / “Staging imaging: no distant metastasis” |
| M1a | Metastasis confined to one distant site or organ, without peritoneal metastasis. *A metastasis in a non-regional lymph node (for example, para-aortic, aortocaval or other retroperitoneal nodes) counts as one distant site.* | Metastases in two or more distant sites or organs; any peritoneal metastasis. | “Two lesions in the right lobe of the liver consistent with metastases; lungs and peritoneum clear.” / “Staging imaging: solitary lung metastasis” |
| M1b | Metastases in two or more distant sites or organs, without peritoneal metastasis. | Metastases confined to one site or organ; any peritoneal metastasis. | “Metastases in the liver and in both lungs; no peritoneal disease.” / “Staging imaging: liver and adrenal metastases” |
| M1c | Metastasis to the peritoneal surface, alone or with metastases in other sites or organs. | No peritoneal metastasis. | “Multiple peritoneal and omental nodules consistent with metastatic carcinoma.” / “Staging imaging: peritoneal carcinomatosis and a liver metastasis” |
| not stated | The report, including any staging imaging, does not say whether distant metastasis is present. | A report whose staging information states whether distant metastasis is present. | “Staging imaging: not available” |

The T4b option gives the AJCC 8th edition definition without its words “or adheres to”; adhesion is dealt with by the note in the second version. The examples of non-regional nodes include “other retroperitoneal nodes”; the pericolic nodes of the ascending and descending colon, which lie in the retroperitoneum, are regional, and no report described such a node.

### Supplementary Note 3. Outcome reference

#### Specification

Data and participants: NHANES 1999–2008 linked to the public-use Linked Mortality File, deaths through 31 December 2019; adults aged 40–79 at examination (Methods; Supplementary Table 9). No eligible participant had an undetermined vital status at 120 months. Outcome: death from any cause within 120 months of examination. Predictors: every field of the record, as the systems saw it; cubic splines at quantile knots for continuous variables, missingness indicators, and interactions of each demographic label with age and of race with sex. Sample size: 11,971 fitting participants, 1,899 deaths (Supplementary Table 9). Missing data: a missing number was set to the fitting-set median with an indicator for that field, and a missing category was kept as a level of its own, in fitting and in prediction alike. 1,856 of 11,971 fitting participants (15.5%) had at least one number set this way, most often income, blood pressure or waist circumference, 61 of them for one or two of the nine biomarkers. Software-test and evaluation participants had all nine. Model building: a fixed light ridge penalty, not tuned; no variable selection; fitted on the fitting set only. Performance: on the evaluation set and on out-of-fold predictions for the fitting set, beside a LightGBM model and a model of age and sex (Supplementary Table 1), and PhenoAge in the first. Uncertainty: 1,000 bootstrap refits of the fitting set (300 for LightGBM), carried into every interval. Code and data: Methods, Code and Data availability.

#### Checks

Participants and data sources: a national sample examined with standardised protocols; the reference is used in the population it was fitted on, without survey weights. Refitted with the survey's examination weights, the reference gave Jev a clinical slope of 0.31. Predictors: measured by protocol at examination, before the outcome and without knowledge of it; the same fields the systems saw. Outcome: death from probabilistic linkage to the National Death Index, ascertained without reference to the predictors; ten-year follow-up complete for every eligible participant. Analysis: 1,899 deaths among 11,971 fitting participants; continuous variables modelled with splines, not categorised; missing values as above; no censoring within 120 months; a fixed ridge penalty against overfitting. On the 1,200 held-out participants the AUROC was 0.826 (95% CI 0.794–0.858) and the calibration slope 0.83 (95% CI 0.72–0.98), close to those of the LightGBM (0.82) and survival (0.89) models on the same records, and 0.93 on the 12,697 records used for comparison (Supplementary Table 1). Effect modification beyond the interactions above was not modelled; the gradient-boosted reference, which models it freely, gave similar clinical slopes (Supplementary Table 10). A discrete-time survival model on the same fields, using follow-up time, gave Jev a clinical slope of 0.39 (simultaneous 95% CI 0.25–0.53). Applicability: the reference describes what outcomes support among US adults aged 40–79 examined in 1999–2008 and followed to 2019. Performance estimates: discrimination and calibration came from ten-fold out-of-fold predictions, with imputation, scaling, spline knots and the fit repeated within each fold, and from held-out participants.

### Supplementary Note 4. Cancer staging: how the reports were built

Except for 18 ordinary reports (last paragraph), the reports were built by code from fixed templates with one random seed (20260922), so each can be rebuilt exactly. Each report stores its T, N and M categories, node counts and number of tumour deposits, so the set can be scored again under another edition.

Stage and categories. The eleven AJCC 8th edition stage groups gave ten levels, IVA and IVB scored as one. For each report, a T, N and M combination was drawn at random from the combinations that define its stage group. Tis occurred only in stage 0, and a stage IVC report that drew T1 or T2 had T redrawn from T3–T4b.

| Category | What the report states |
|---|---|
| N0, N1c | No involved regional node |
| N1a / N1b / N2a / N2b | 1 / 2–3 / 4–6 / 7–14 involved nodes |
| Nodes examined | At least 12, and at least 3 more than the involved count, plus a right-skewed extra (mean 8) |
| Tumour deposits | 1–4 in every N1c report and in 25% of other node-positive reports |
| M0 | Staging computed tomography: no distant metastasis |
| M1a / M1b | Metastasis in one organ (liver or lung) / in two (liver, lung or bone) |
| M1c | Peritoneal disease, with or without an organ metastasis |
| T4b | Direct invasion of small bowel or the abdominal wall |

Findings that do not change the stage were drawn at fixed rates: site (seven colon segments, each with its operation), mucinous histology 15%, high grade 20%, lymphovascular invasion 30%, perineural invasion 20%, deficient mismatch repair 15%, negative margins (closest 0.5–8.0 cm), and tumour size and thickness within ranges that grow with T (for example 0.8–3.0 cm for Tis and 2.5–9.0 cm for T3). No report described neoadjuvant therapy, and no ordinary report described isolated tumour cells or an adhesion without tumour.

Wording. The report follows the College of American Pathologists colon resection case summary, in its synoptic layout or as narrative, with distant metastasis stated in a staging computed tomography line under the clinical information. No report states a T, N or M category or a stage. Each finding has three phrasings per category and layout, chosen at random, and the first phrasing of each depth category follows the AJCC definition. Report identifiers were assigned in random order.

Checks. A unit test covers every cell of the AJCC table (144 cells). A parser written separately from the templates recovered the intended categories and counts from every template-built report. No two reports are identical, including after normalising whitespace and case and masking every number. A physician author reviewed every phrase that encodes a category. The 100 reports with a misleading feature and the 20 with a node at an unusual site were built by the same code, with the category combination fixed.

Eighteen ordinary reports came from a second set, with categories drawn from the same staging table (seed 20260928) and wording varied by a language model outside the study; none contained a TNM code or the words stage, AJCC, TNM, Dukes or UICC, and two independent model readers recovered the intended categories from each.

3. World Health Organization. Obesity: preventing and managing the global epidemic. WHO Technical Report Series 894. World Health Organization, 2000.
